# Polymethylation Scores for Prenatal Maternal Smoke Exposure Persist Until Age 15 and Are Detected in Saliva in the Fragile Families and Child Wellbeing Cohort

**DOI:** 10.1101/2021.11.30.21267020

**Authors:** Freida A. Blostein, Jonah Fisher, John Dou, Lisa Schneper, Erin B. Ware, Daniel A. Notterman, Colter Mitchell, Kelly M. Bakulski

## Abstract

Prenatal maternal smoking is associated with low birthweight, neurological disorders, and asthma in exposed children. DNA methylation signatures can function as biomarkers of prenatal smoke exposure. However, the robustness of these DNA methylation signatures across child ages, genetic ancestry groups, or tissues is not clear. Using coefficients from a meta-analysis of prenatal maternal smoke exposure and DNA methylation in newborn cord blood, we created polymethylation scores of saliva DNA methylation from children at ages 9 and 15 in the Fragile Families and Child Wellbeing study. In the full sample at age 9 (n=753), prenatal maternal smoke exposure was associated with a 0.51 (95%CI: 0.35, 0.66) standard deviation higher polymethylation score. The direction and magnitude of the association was consistent in European and African genetic ancestry samples. In the full sample at age 15 (n=747), prenatal maternal smoke exposure was associated with a 0.48 (95%CI: 0.32, 0.63) standard deviation higher polymethylation score, and the association was attenuated among the European and Admixed - Latin genetic ancestry samples. The polymethylation score classified prenatal maternal smoke exposure accurately (AUC age 9=0.77, age 15=0.76). Including the polymethylation score increased the AUC of base model covariates by 5 (95% CI: (2.1, 7.2)) percentage points, while including a single candidate site in the *AHRR* gene did not (*P*-value=0.19). Polymethylation scores for prenatal maternal smoking were portable across genetic ancestries and more accurate than an individual DNA methylation site. DNA polymethylation scores from saliva samples could serve as robust and practical clinical biomarkers of prenatal maternal smoke exposure.

## Introduction

*In utero* cigarette smoke exposure causes low birth weight and preterm birth [1]. Exposure to cigarette smoking *in utero* is also associated with a variety of short- and long-term postnatal adverse health outcomes [2]. These include conditions with life-long consequences, including negative neurodevelopmental outcomes, obesity, and asthma [3]. Although 11% of US pregnant people in 2018 reported any past-month cigarette use [4], this is considered an underestimate. When compared to biomarker measures, maternal prenatal smoking is underreported - presumably due to stigma [5, 6]. The gold-standard smoking biomarker, serum cotinine levels, has a half-life of nine hours in pregnant people, and is rarely available outside of birth cohort research [7]. Alternative biomarkers of prenatal maternal smoke exposure with a larger window of measurement could improve measurement of prenatal smoke exposure in older children and adults.

DNA methylation is a possible biomarker of prenatal smoke exposure. Smoking during pregnancy is associated with offspring DNA methylation signatures at birth in placental and cord blood samples, and postnatally in peripheral blood samples [8–13]. Another postnatal biospecimen option is saliva, which is easier to collect than blood and increasingly collected in large epidemiological cohorts [14]. However, associations of pregnancy smoking with offspring salivary DNA methylation have not been measured. Since DNA methylation contributes to cell differentiation and cell type proportions differ across tissue types, it is important to verify associations in new tissues [15]. In addition, most studies of prenatal maternal smoking and childhood DNA methylation have been conducted in primarily European genetic ancestry populations [9, 13, 16–19], and it will be important to test generalizability to other groups. Few studies have assessed multiple genetic ancestry groups in the same cohort and no study has evaluated associations between pregnancy cigarette smoking and salivary DNA methylation in children.

Using the findings from large-scale association testing in independent populations, DNA methylation-based exposure biomarkers can be developed and assessed by evaluating their classification accuracy. Biomarkers can range from single DNA methylation sites to summary measures across multiple sites, termed polymethylation scores [22]. In a polymethylation score, DNA methylation sites are weighted by their effect size from a previously conducted analysis in an independent sample and then summed to a single score [23, 24]. DNA polymethylation scores for *in utero* smoke exposure have performed well (area under the curves (AUCs)>0.8) in cord blood samples from newborns and peripheral blood samples from young children [16, 21]. Polymethylation scores have also classified *in utero* smoke exposure when using from DNA methylation in peripheral blood from middle-aged adults (AUC of 0.72 (95% confidence interval 0.69, 0.76)) [12]. Yet few studies have assessed the longitudinal persistence of DNA-methylation-based smoking biomarker accuracy.

DNA methylation differences hold promise as an exposure biomarker for *in utero* cigarette smoke. We must understand how the signal varies across tissue, genetic ancestry, and age. In the Fragile Families and Child Wellbeing Study, a diverse longitudinal birth cohort, we aimed to assess salivary DNA methylation biomarkers for pregnancy cigarette smoking. We tested associations between pregnancy smoke exposure and DNA methylation, measured at ages 9 and 15. We compared the magnitude and specificity of these signals to other DNA methylation measures including individual a priori DNA methylation sites, global DNA methylation, and epigenetic clocks. We then performed genetic ancestry- and age-stratified analyses to test the hypothesis that DNA methylation could serve as a portable and persistent biomarker of prenatal maternal smoking.

## Methods

### Cohort

The Fragile Families and Child Wellbeing Study is a birth cohort of nearly 5,000 children born in 20 cities in the United States between 1998 and 2000 [25]. Participants were selected at delivery using a three-stage stratified random sample design which oversampled unmarried mothers by a ratio of 3:1 [25]. Participants were excluded at the baseline wave on the following criteria: the parents who planned to place the child for adoption, the father was deceased, the parents did not speak English or Spanish well enough to be interviewed, the mother or baby were too ill to complete the interview, or the baby died before the interview could take place. Children were followed longitudinally with assessments at ages 1, 3, 5, 9, and 15; at age 22, follow up is ongoing. Across certain child ages, assessments included medical record extraction, biosample collection, in-home assessments, and surveys of the mother, father, primary caregiver, teacher, and child. At ages 9 and 15, a saliva sample was taken from the child [25]. A subsample of the Fragile Families cohort was selected for saliva DNA and DNA methylation processing [25]. To be eligible for the DNA methylation assessment, children had to have participated and given saliva at both ages 9 and 15, after which a random sample of these eligible participants was then selected. One exception to this, all participants from Detroit, Toledo, and Chicago (participants in the sub-Study of Adolescent Neurodevelopment (SAND) [26]) were assayed for DNA methylation, even if they only provided a sample at one time-point. All analyses account for oversampling of these participants.

### Pre- and postnatal exposure measurement

At time of child’s birth, mothers answered categorical questions about their pregnancy smoking, drug, and alcohol use. For maternal prenatal smoking, mothers were asked: “During your pregnancy, how many cigarettes did you smoke?” Responses were categorical: 2 or more packs a day, 1 or more but less than 2 packs per day, less than 1 pack a day, or none. Few participants reported smoking a pack or more a day. Thus, for our primary exposure variable, we created a dichotomous (any versus no prenatal smoke exposure) variable.

Mothers were also asked how frequently they drank during their pregnancy. We created a dichotomous any versus no prenatal alcohol exposure variable. Similarly, mothers were asked how frequently they used any other drug during their pregnancy. We again created a binary variable (any versus no prenatal use of other drugs).

When the children were 1, 5, 9, and 15 years of age, primary caregivers responded to questions about smoking in the household. To encapsulate early life postnatal secondhand smoke exposure, we created a binary variable for caregiver smoking behavior at ages 1 and/or 5 (any versus no). To encapsulate recent postnatal smoke exposure to the age 9 and 15 interview where DNA methylation was collected, we used a caregiver categorical variable for packs per day (no smoking, less than one pack/day, one or more packs/day) in the month prior to each interview.

At age 9 children were asked if they had ever smoked a cigarette or used tobacco (yes/no), and at age 15 they were asked if they had ever smoked an entire cigarette (yes/no).

### Covariate measurement

We used household income-to-poverty ratio at baseline as a proxy for socioeconomic status. Income-to-poverty ratio was the ratio of total household income (as self-reported by the mother) to the official poverty thresholds designated by the United States Census Bureau for the year preceding the interview.

Mothers reported sex (male/female) of their child at baseline.

Child genetic ancestry was calculated. Principal components (PC) of child genetic ancestry were calculated from genetic data measured using PsychChip (Illumina, San Diego, California) with child saliva samples [27]. Genetic ancestry was assigned by comparing PC loadings to 1000 Genomes super population clusters. Samples with PC1 > 0.018 and PC2 > -0.0075 were assigned to European ancestry. Samples with PC1 < - 0.005 and PC2 > 0.007+0.75(PC1) were assigned to African ancestry. Samples with PC1 > 0.018 and -0.055 < PC2 < 0.025 were assigned to Admixed ancestry [27]. Samples assigned to Admixed ancestry were primarily from individuals whose mothers self-reported Hispanic ethnicity (76% of samples), so we label this group Admixed - Latin ancestry.

### DNA methylation measurement

Salivary samples from the children were collected at ages 9 and 15 using the Oragene DNA sample collection kit (DNA Genotek Inc., Ontario). Saliva DNA was extracted manually using DNA Genotek’s purification protocol using prepIT L2P. DNA was bisulfite treated and cleaned using the EZ DNA Methylation kit (Zymo Research, California). Age 9 and 15 samples were run on the same slide to minimize technical variation, and otherwise samples were randomized. Saliva DNA methylation was measured using the Illumina HumanMethylation 450k BeadArray [28] and imaged using the Illumina iScan system. The bead arrays were processed at the Genome Sciences Core Facility at the Pennsylvania University College of Medicine (Hershey, PA).

DNA methylation image data were processed in R statistical software (4.1) using the Enmix package [29]. We used the February 2022 data freeze of the Fragile Families and Child Wellbeing DNA methylation data. The red and green image pairs (n=1,811) were read into R and the ENmix *preprocessENmix* and *rcp* functions were used to normalize dye bias, apply background correction and adjust for probe-type bias. Further quality control was applied using the ewastools packages [30]. We dropped samples using the following criteria: if >10% of DNA methylation sites had detection p-value >0.01 or a beadcount under 4 (n=34), if there was sex discordance between DNA methylation predicted sex and recorded sex (n=11), if samples had outlier methylation values (n=6) or if two sequential samples from the same individual exhibited genetic discordance between visits (n=27) (Figure 1). Technical replicates were removed (n=49). DNA methylation sites were removed if they had detection p-value >0.01 or a beadcount under 4 in ≥ 5% of samples (n=47,930) [31]. Relative proportions of immune and epithelial cell types were estimated from DNA methylation measures using a childhood saliva reference panel [32].

**Figure 1.**
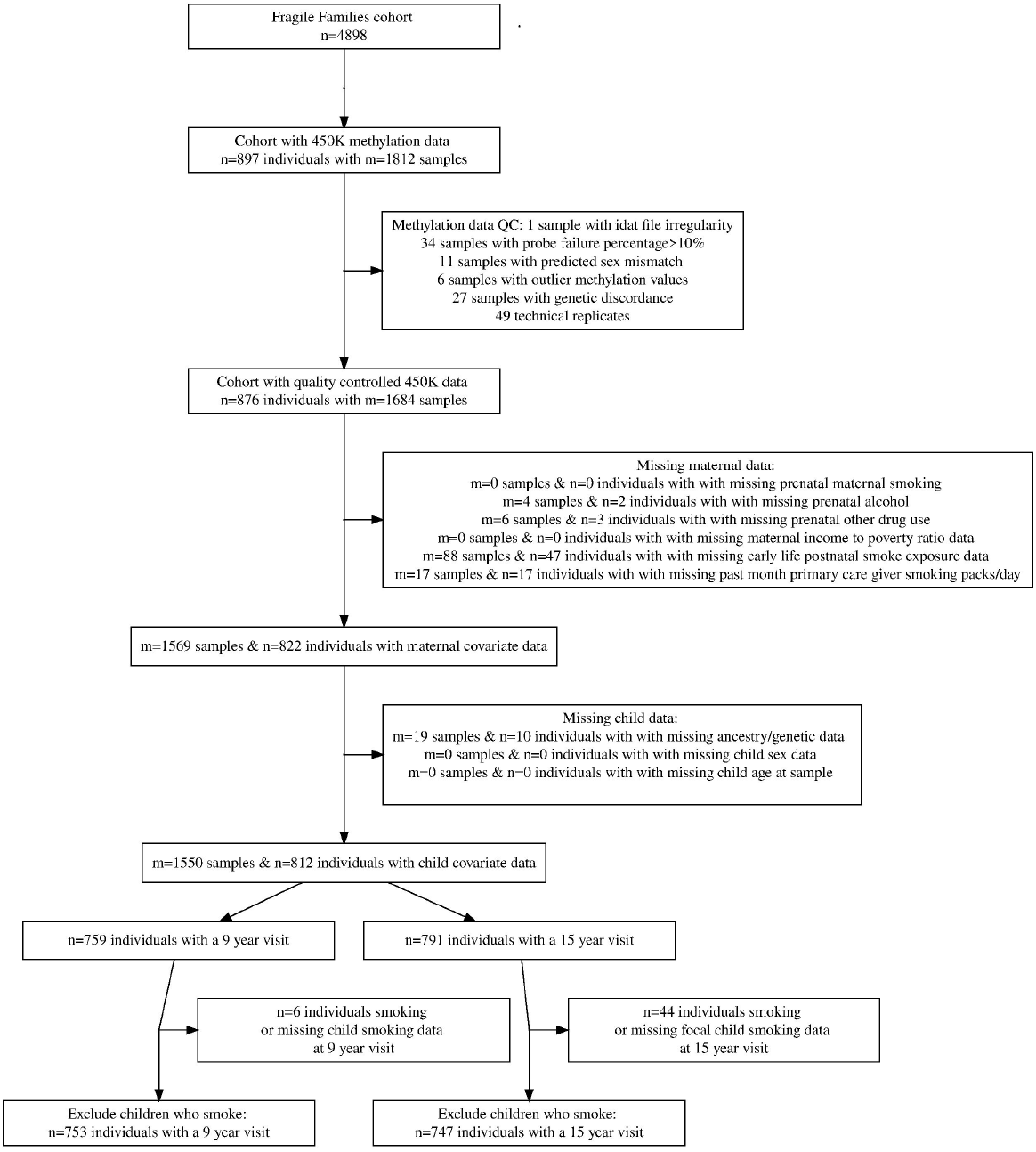
Selection of samples from the Fragile Families and Child Wellbeing study into analytic subset. N represents the number of individuals at each step in the selection procedure, M represents the number of samples. Individuals with repeated measures can have more than one sample.

Our primary outcome variable was a polymethylation score for prenatal maternal smoke exposure. From an independent meta-analysis of prenatal smoke exposure and DNA methylation (the 2016 Joubert meta-analysis), we extracted the regression coefficients of 6,073 DNA methylation sites associated with prenatal maternal smoking at a false discovery ratio corrected *P* value <0.05 [9]. Of the 6,073 CpG sites with regression coefficients, 5,666 were available in our dataset after quality filtering (Supplemental Table 1). For our main analysis we used coefficients from a regression of sustained smoking exposure and DNA methylation in newborn cord blood with cell-type control. We mean-centered the DNA methylation beta values in our study, weighted them by the independent regression coefficients, and took the sum across sites. This resulted in a single smoking polymethylation score per sample. To facilitate interpretation, we z-score standardized the polymethylation scores.

We investigated several sensitivity measures of DNA methylation, including several alternative polymethylation scores for smoke exposure, single DNA methylation sites, mean global DNA methylation, and DNA methylation clocks.

To assess the robustness of polymethylation scores to the source of coefficients for weights, we generated two sensitivity polymethylation scores from the 2016 Joubert meta-analysis [9]. First, we used the regression of cord blood DNA methylation sites associated with prenatal maternal smoking at a strict Bonferroni threshold (n=569 sites, 549 sites available in our data), when analyses were not adjusted for cell proportions [9]. Second, we used the regression coefficients of child peripheral blood DNA methylation sites associated with prenatal maternal smoking at the strict Bonferroni threshold (n=19 sites, 19 sites available in our data) when analyses were not adjusted for cell proportions [9].

To assess the robustness of polymethylation scores to the source and selection of DNA methylation sites, we generated two additional sensitivity polymethylation scores. One score was generated using DNA methylation sites and coefficients prioritized from a LASSO regression of prenatal maternal smoking and cord blood methylation in the Norwegian Mother and Child Cohort Study (n=28 sites, 27 sites available in our data) [21]. Another score was generated using DNA methylation sites and coefficients prioritized from an elastic net regression of prenatal maternal smoking and DNA methylation in adolescent peripheral blood (n=204 sites, 191 sites available in our data) [33]. This score was calculated using the coefficients and code provided by the authors in the R code accompanying the manuscript [34]. We provide a table detailing the exact CpG sites and coefficients available for each score used in this analysis (Supplemental Table 1).

To compare the utility of polymethylation scores relative to single site biomarkers, we selected the top DNA methylation site from the 2016 Joubert meta-analysis, *AHRR:* cg05575921. As a sensitivity analyses, we also selected four other *a priori* DNA methylation sites which were identified from meta-analyses of prenatal maternal smoking and DNA methylation in children’s cord and peripheral blood samples: *GFI1*: cg14179389, *CYP1A1*: cg05549655, *MYO1G*: cg22132788, and *MYO1G*: cg04180046 [9, 35].

To assess the specificity of the impact of prenatal smoke exposure on DNA methylation, we compared the polymethylation scores to mean global and regulatory regional DNA methylation and DNA methylation-derived age measures. Global DNA methylation was calculated for each sample as the mean methylation value of each sample across the pre-processed array. Mean DNA methylation for each genomic regions (CpG island, shore, shelf or open sea, as identified in the R package IlluminaHumanMethylation450kanno.ilmn12.hg19 v 0.6.0) was also calculated. Pediatric epigenetic age was calculated for each sample using the coefficients and methods provided by the creators (see <https://github.com/kobor-lab/Public-Scripts>) [36]. As a sensitivity analysis, the GRIM-age clock, which was neither trained nor validated in a pediatric population, was calculated as previously described [37].

To control for unmeasured confounding and potential technical variation, we calculated surrogate variables from DNA methylation data using the sva R package (version 3.38) [38]. The relationship between prenatal maternal smoking and DNA methylation was protected. To describe variability in the DNA methylation data, we evaluated the association between surrogate variables and technical covariates (sample plate, slide, and position on slide) using a heatmap of P values from appropriate statistical tests.

### Statistical analyses

#### Inclusion exclusion criteria

Among samples with pre-processed DNA methylation values, we excluded samples missing any data on the following variables: maternal prenatal smoking, maternal prenatal alcohol use, maternal prenatal other drug use, maternal income to poverty ratio, postnatal secondhand smoke exposure variables, child sex, or child age. We also excluded samples that were missing genetic data. To ensure we measured the direct effect of prenatal exposure to cigarette smoke, rather than the effect of personal postnatal smoking, we also excluded children who reported own-smoking. Specifically, samples from children who reported ever smoking a cigarette or using tobacco at age 9 or were missing own-smoking data were excluded. Samples from children who reported smoking a cigarette at age 9 or at age 15 or were missing own-smoking data were excluded. Children who were missing a response to the question at age 9 but answered that they had never smoked a whole cigarette at age 15 were kept in the sample. Sample inclusion was visualized using a flow chart.

#### Descriptive statistics

We described the distribution of continuous variables using the mean and standard deviation. We described the distribution of categorical variables using frequencies. We compared the distribution of included to excluded samples using the Welch’s t-test for continuous variables and the Chi-Square test for categorical variables. We reported the correlation between continuous measures of DNA methylation (including polymethylation scores, single sites, mean global DNA methylation and estimated cell type proportions) using pairwise Pearson correlation coefficients. We reported the correlation of these DNA methylation measures between visits using pairwise Pearson correlation coefficients and scatter plots. We compared the distribution of variables by exposure status to prenatal maternal smoking using the Welch’s t-test for continuous variables and the Chi-Square test for categorical variables. For select continuous DNA methylation measures we compared the distributions across prenatal smoke exposure status using violin plots with captive box plots.

#### Regression analyses

To assess the association between our primary exposure variable (prenatal maternal smoking) and primary outcome variable (salivary DNA methylation polymethylation score), we performed age-stratified, multivariable linear regressions. We adjusted for an indicator variable for maternal residence in one of the oversampled cities (Chicago, Detroit, or Toledo), maternal income to poverty ratio at baseline, child sex, child age, plate from DNA methylation processing (approximately 96 samples per plate), estimated immune cell proportion from DNA methylation data, and the first two components of genetic ancestry from principal component analysis. While children provided saliva samples at two visits corresponding to ages 9 and 15, there was variation in the children’s age in months at each visit. To account for this potential variability, age of the child in months was controlled for within age-stratified models. While child sex, child age, plate from DNA methylation processing, and immune cell proportions are not confounders (as they cannot causally affect prenatal maternal smoke exposure), these variables can strongly affect DNA methylation and so were adjusted for as precision variables. We reported the effect estimate for polymethylation score term and the 95% confidence interval (CI). We visualized our findings using forest plots.

To assess the portability of the association between our primary exposure variable of prenatal maternal smoking and salivary DNA methylation across genetic ancestry, we additionally stratified by genetic ancestry and reran the models described above. In ancestry-stratified models, the first two principal components from principal component analysis run within each ancestry strata were used.

To compare the utility of the polymethylation score to a single site biomarker, we used percent methylation at AHRR: cg05575921 as the outcome in the models described above. To assess the specificity of the impact of prenatal maternal smoking on DNA methylation, we used mean percent global DNA methylation and the pediatric DNA methylation clock as outcome variables.

#### Regression sensitivity analyses

To assess the specificity of the DNA methylation signal to the type and timing of exposure, we additionally adjusted for the following variables in sensitivity analyses: 1) prenatal drug and alcohol use and 2) postnatal second-hand smoke exposure in addition to prenatal drug and alcohol use. To assess the robustness of the association between prenatal maternal smoking and DNA methylation to technical variation and residual confounding, we also regressed DNA methylation against prenatal maternal smoking exposure while adjusting for surrogate variables calculated from the DNA methylation data.

To assess the robustness of the association to the choice of polymethylation score, we performed the same regression analyses described above with sensitivity polymethylation scores as the outcome. Similarly, we replaced the outcome variable of *AHRR:* cg05575921 with other *a priori* selected DNA methylation sites in sensitivity analyses. We plotted the estimates and 95% CI for each DNA methylation site along with the previously reported estimates from the literature using forest plots [9, 35]

#### Receiver operator curve analysis

To evaluate the accuracy of the DNA methylation summary measures as biomarkers of prenatal maternal smoking, we used a receiver operating curve [39]. First, we regressed exposure to prenatal maternal smoking (outcome) against the DNA methylation summary measures in individual logistic regressions, while adjusting for the base model variables from regression analyses. The base model variables included indicator variable for maternal residence in one of the oversampled cities (Chicago, Detroit or Toledo), maternal income to poverty ratio at baseline, child sex, child age, plate from DNA methylation processing, estimated immune cell proportion from DNA methylation data, and the first two components of genetic ancestry from principal component analysis. Next, we calculated receiver operating curves (ROC) and area under the curves (AUCs) using the function *roc* from the R library pROC version 1.18.0 [40]. We compared ROC curves and AUCs from models with only base model variables to those which included base model variables and the methylation summary measure using the Delong method and the function *roc*.*test* from the pROC library. This is appropriate as the base model is nested within the base model variable + methylation summary measures model.

#### Receiver operator curve sensitivity analyses

To assess the robustness of polymethylation scores classification accuracy to the choice of polymethylation score coefficients, we used polymethylation scores calculated using different coefficient sets as predictors for the receiver operating curve analysis in sensitivity analyses.

To compare the performance of saliva-based polymethylation scores to previous reports of blood-based polymethylation scores, we used a forest plot. We plotted the AUC and 95% CI (when available) for each polymethylation score in our cohort and in the cohorts previously reported in the literature [12, 33].

## Results

### Study sample descriptive statistics

Among participants in the Fragile Families and Child Wellbeing Study, saliva DNA methylation was measured on 1,812 samples from 897 unique child participants with the Illumina 450K array. Complete data on covariates of interest was available on 1,500 samples from 805 participants who were included in the analysis (Figure 1). There were 695 participants with both age 9 and age 15 DNA methylation samples. Excluded participants were similar to included participants, except that included participants were slightly more likely to be of African genetic ancestry and less likely to be of Admixed-Latin genetic ancestry (Supplemental Table 2). In the analytic sample, 20% percent of the mothers reported any prenatal maternal smoking, 12% reported prenatal alcohol use, and 5% reported prenatal drug use. The mean income to poverty ratio of mothers at birth was 2.22. Of the included children, 50% were male, 61% were of African genetic ancestry, and 24% were of Admixed-Latin genetic ancestry.

All of the polymethylation scores for prenatal smoke exposure were correlated with each other (Supplemental Figure 1). Our primary polymethylation score for prenatal smoke exposure, which was calculated using coefficients from a cell-type corrected regression, was weakly correlated with estimated cell-type proportion of immune cells (Pearson *r*=0.11; *P*-value <0.001).

Among the 695 individuals with data from both the ages 9 and 15 visits, we compared epigenetic measures over time (Supplemental Figure 2). The correlation across ages was strongest for the polymethylation scores for prenatal smoke exposure constructed using the 2016 Joubert meta-analysis coefficients from newborn cord blood (Pearson *r*=0.91, *P*-value<0.0001). Global DNA methylation (*r*=0.68, *P*-value<0.0001) and epigenetic clocks were less strongly correlated across ages (Pediatric: r=0.49; *P*-value<0.0001, GRIM: *r*=0.49; *P*-value<0.0001) Epigenetic ages were higher at the age 15 visit than the age 9 visit, as expected (Supplemental Figure 3).The distributions of global methylation and polymethylation scores were more consistent (Supplemental Figure 3). Almost all saliva samples consisted of primarily immune cells, based on estimated cell-type proportions. At age 9, the mean estimated immune cell proportion was 96% (minimum=25%, first quartile=99%, third quartile=100%, maximum=100%). At age 15, the mean estimated immune cell proportion was 93 % (minimum=3%, first quartile=92%, third quartile=100%, maximum=100%).

### Bivariate associations between prenatal maternal smoking and DNA methylation summary measures

Mothers who reported smoking during pregnancy had lower income to poverty ratios (mean=1.5) than those who did not (mean=2.4). Mothers who smoked were more likely to report prenatal alcohol use (30% vs 7%), prenatal drug use (18% vs 2%), and postnatal smoking (96% vs 25%) than mothers who did not report prenatal maternal smoking (Table 1). Children of mothers who reported smoking during pregnancy were more likely to be of European genetic ancestry (62%) than children of mothers who did not (60%, *P*-value=<0.001). Children of mothers who reported smoking during pregnancy had higher prenatal maternal smoking polymethylation scores than children of mothers who did not at both age 9 (0.1 vs -0.04, *P*-value<0.001) and age 15 (0.13 vs -0.01, *P*-value<0.001) visits (Table 1, Supplemental Table 3). At age 9, children exposed to prenatal maternal smoking had lower DNA methylation at cg05575921 (76.7%) than children of non-smoking mothers (77.6%, *P*-value=0.07). At age 15, children exposed to prenatal smoking had lower DNA methylation at cg05575921 (75.8%) than children of non-smoking mothers (76.5% *P*-value= 0.22). Epigenetic age from the pediatric and GRIM clocks and global DNA methylation did not differ between children exposed versus unexposed to prenatal maternal smoking (Figure 2).

**Table 1:** Bivariate associations between prenatal maternal smoking (no/yes) and selected DNA methylation summary measures and important covariates among a diverse sample of 805 children in the Fragile Families and Child Wellbeing study

| Characteristic | Age 9 |  |  |  | Age 15 |  |  |  |
| --- | --- | --- | --- | --- | --- | --- | --- | --- |
|  | N | Not exposed,<br>N = 598<sup>1</sup> | Smoke exposed,<br>N = 155<sup>1</sup> | p-value<sup>2</sup> | N | Not exposed,<br>N = 604<sup>1</sup> | Smoke exposed,<br>N = 143<sup>1</sup> | p-value<sup>2</sup> |
| Child characteristics |  |  |  |  |  |  |  |  |
| Polymethylation score for smoke exposure (coefficients from 2016 Joubert meta-analysis of newborn cord blood with cell-type control, 6073 DNA methylation sites) | 753 | -0.04<br>(0.24) | 0.10<br>(0.25) | <0.001 | 747 | -0.01<br>(0.25) | 0.13<br>(0.25) | <0.001 |
| AHRR gene: percent cg05575921 methylation | 753 | 77.6<br>(5.4) | 76.7<br>(5.6) | 0.070 | 747 | 77 (6) | 76 (6) | 0.2 |
| Pediatric epigenetic clock (years) | 753 | 9.80<br>(1.07) | 9.70<br>(1.13) | 0.3 | 747 | 13.27<br>(2.64) | 13.24<br>(2.57) | 0.9 |
| Percent global DNA methylation | 753 | 50.32<br>(0.95) | 50.34<br>(1.08) | 0.9 | 747 | 50.19<br>(1.07) | 50.28<br>(1.01) | 0.3 |
| Immune cell proportion (saliva) | 753 | 0.96<br>(0.10) | 0.97<br>(0.08) | 0.3 | 747 | 0.93<br>(0.15) | 0.93<br>(0.14) | 0.9 |
| Epithelial cell proportion (saliva) | 753 | 0.04<br>(0.10) | 0.03<br>(0.08) | 0.3 | 747 | 0.07<br>(0.15) | 0.07<br>(0.14) | 0.9 |
| Ancestry categorization from child principal components of genetic data | 753 |  |  | 0.001 | 747 |  |  | 0.002 |
|  | N | Not exposed,<br>N = 598<sup>1</sup> | Smoke exposed,<br>N = 155<sup>1</sup> | p-value<sup>2</sup> | N | Not exposed,<br>N = 604<sup>1</sup> | Smoke exposed,<br>N = 143<sup>1</sup> | p-value<sup>2</sup> |
| Admixed ancestry<br>- Latin heritage |  | 157<br>(26%) | 23<br>(15%) |  |  | 150<br>(25%) | 21<br>(15%) |  |
| African ancestry |  | 358<br>(60%) | 96<br>(62%) |  |  | 369<br>(61%) | 88<br>(62%) |  |
| European ancestry |  | 83<br>(14%) | 36<br>(23%) |  |  | 85<br>(14%) | 34<br>(24%) |  |
| Child gender | 753 |  |  | 0.8 | 747 |  |  | 0.9 |
| Boy |  | 301<br>(50%) | 76<br>(49%) |  |  | 295<br>(49%) | 69<br>(48%) |  |
| Girl |  | 297<br>(50%) | 79<br>(51%) |  |  | 309<br>(51%) | 74<br>(52%) |  |
| Maternal characteristics |  |  |  |  |  |  |  |  |
| Oversampled cities | 753 |  |  | 0.13 | 747 |  |  | 0.6 |
| Detroit, Chicago<br>or Toledo |  | 149<br>(25%) | 48<br>(31%) |  |  | 175<br>(29%) | 45<br>(31%) |  |
| Not Detroit,<br>Chicago or Toledo |  | 449<br>(75%) | 107<br>(69%) |  |  | 429<br>(71%) | 98<br>(69%) |  |
| Maternal prenatal<br>alcohol use | 753 | 44<br>(7.4%) | 47<br>(30%) | <0.001 | 747 | 40<br>(6.6%) | 43<br>(30%) | <0.001 |
| Maternal prenatal<br>any drug use | 753 | 10<br>(1.7%) | 26<br>(17%) | <0.001 | 747 | 9<br>(1.5%) | 28<br>(20%) | <0.001 |
|  | N | Not exposed,<br>N = 598<sup>1</sup> | Smoke exposed,<br>N = 155<sup>1</sup> | p-value<sup>2</sup> | N | Not exposed,<br>N = 604<sup>1</sup> | Smoke exposed,<br>N = 143<sup>1</sup> | p-value<sup>2</sup> |
| Maternal income to poverty threshold (at baseline) | 753 | 2.42<br>(2.60) | 1.53<br>(1.56) | <0.001 | 747 | 2.43<br>(2.60) | 1.53<br>(1.64) | <0.001 |
| Postnatal maternal smoking at ages 1 or 5 | 753 |  |  | <0.001 | 747 |  |  | <0.001 |
| No maternal smoking at age 1 and 5 |  | 448<br>(75%) | 7<br>(4.5%) |  |  | 450<br>(75%) | 7<br>(4.9%) |  |
| Maternal smoking at age 1 or age 5 |  | 150<br>(25%) | 148<br>(95%) |  |  | 154<br>(25%) | 136<br>(95%) |  |
| Postnatal primary caregiver smoking in past month prior to visit | 753 |  |  | <0.001 | 747 |  |  | <0.001 |
| No smoking |  | 476<br>(80%) | 23<br>(15%) |  |  | 511<br>(85%) | 42<br>(29%) |  |
| Less than pack a day |  | 95<br>(16%) | 85<br>(55%) |  |  | 81<br>(13%) | 78<br>(55%) |  |
| Pack or more a day |  | 27<br>(4.5%) | 47<br>(30%) |  |  | 12<br>(2.0%) | 23<br>(16%) |  |

<sup>1</sup> Mean (SD); n (%)
<sup>2</sup> Welch Two Sample t-test; Pearson's Chi-squared test

**Figure 2.**
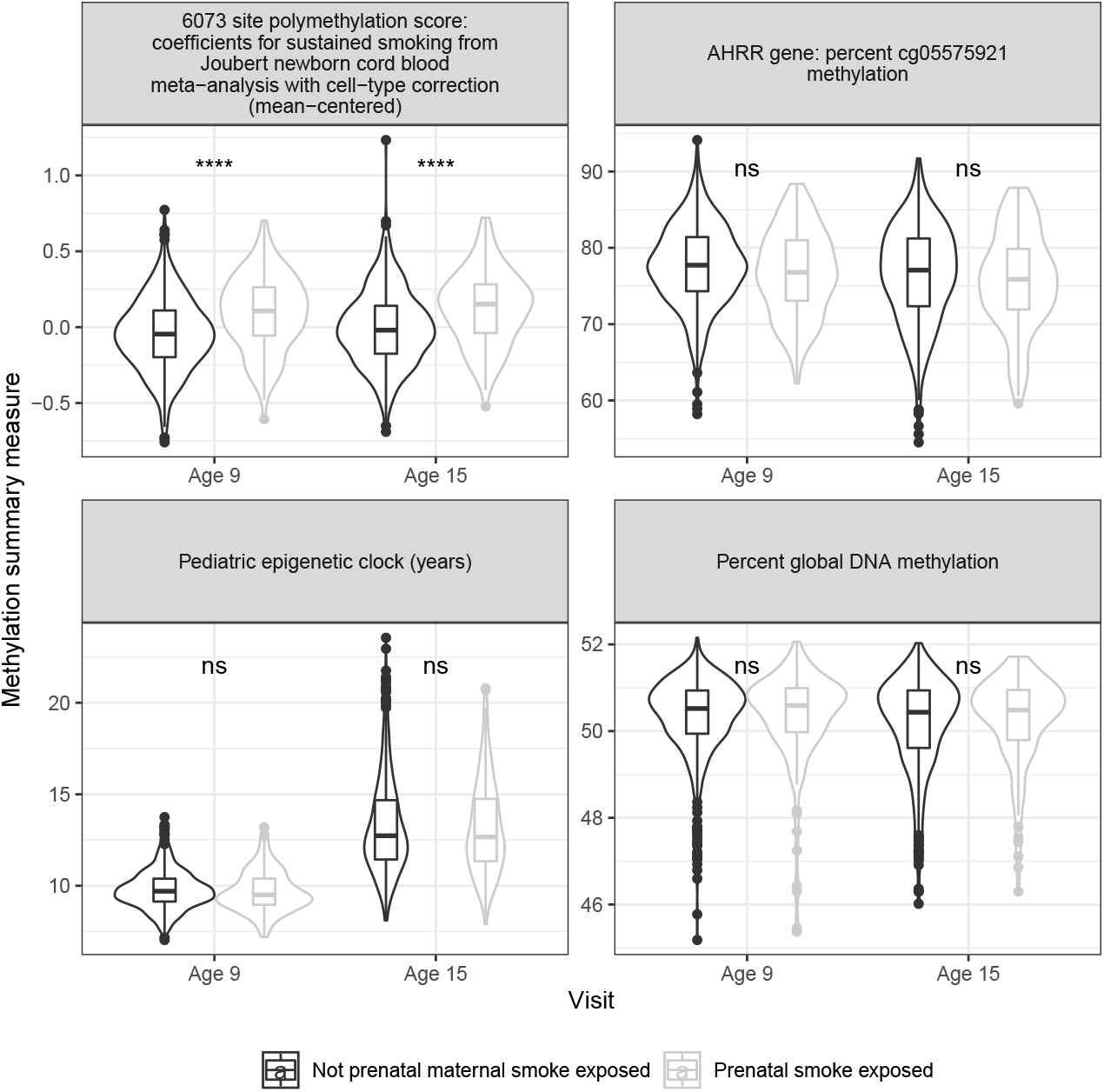
Differences in selected DNA methylation summary measures by self-report of prenatal maternal smoking among 805 children in the Fragile Families and Child Wellbeing study at ages 9 and 15. Samples from children exposed to prenatal maternal smoke in grey, samples from children unexposed to prenatal maternal smoke in black. From top-left, clockwise: Polymethylation scores for prenatal maternal smoke exposure, constructed using regression coefficients for prenatal smoke exposure predicting DNA methylation in newborn cordblood samples, accounting for cell-type control. DNA methylation values from samples in the Fragile Families and ChildWellbeing study were mean-centered, then multiplied by these regression coefficients and summed. Pediatric epigentic clock (years). AHRR gene: percent cg05575921 methylation. Percent global DNA methylation.

### Multivariable associations between prenatal maternal smoking and DNA methylation summary measures

The association between prenatal maternal smoking and the polymethylation score for prenatal smoke exposure was also observed in multivariable models, adjusting for base model covariates of city indicator, child sex, maternal income to poverty ratio at baseline, proportion of salivary immune cells, sample plate from DNA methylation analysis, child age, and the first two principal components of genetic ancestry (Figure 3; Supplemental Table 4). At age 9, prenatal maternal smoke exposure was associated with 0.51 (95%CI: 0.35, 0.66) standard deviation higher polymethylation score for prenatal smoke exposure. At age 15, prenatal maternal smoke exposure was associated with 0.48 (95%CI: 0.32, 0.63) standard deviation higher prenatal smoke exposure polymethylation score for prenatal smoke exposure.

**Figure 3.**
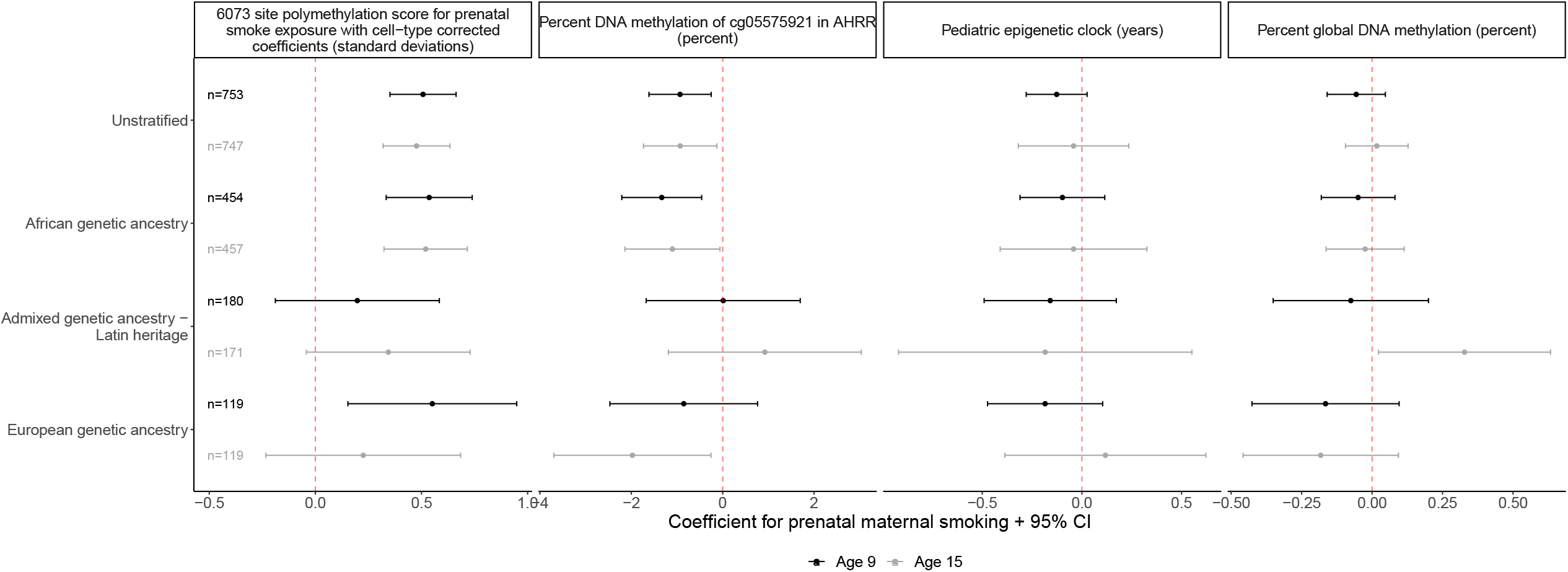
Prenatal maternal smoke exposure is consistently associated with polymethylation scores at ages 9 and 15 and is portable across genetic ancestry groups in a sample of 805 children in the Fragile Families and Child Wellbeing study. All models shown controlled for: first two principal components of child genetic ancestry (from ancestry-stratified principal components for ancestry stratified models), child sex, maternal income-to-poverty ratio at birth, immune cell proportion estimated from methylation data, yes/no other maternal prenatal drug use, yes/no maternal prenatal alcohol use, postnatal maternal smoking when child age 1 or age 5, postnatal maternal/primary care give smoking packs/day in month prior to saliva sample.

A consistent association was observed when stratifying by genetic ancestry. In the African genetic ancestry sample (n= 488) at age 9, prenatal maternal smoking was associated with 0.54 (95%CI: 0.33, 0.74) standard deviation higher polymethylation score for prenatal smoke exposure. The effect was similar among European ancestry children at age 9 (0.55 (95%CI: 0.15, 0.95)) and consistent in direction but attenuated at age 15 (0.23 (95%CI: -0.23, 0.68)). The association was attenuated among Admixed - Latin ancestry children at ages 9 (0.2 (95%CI: -0.19, 0.58)) and 15 (0.34 (95%CI: -0.04, 0.73)) (Figure 3; Supplemental Table 4).

At age 9, prenatal maternal smoking was associated with 0.93 percentage point lower DNA methylation at cg055975921 (95% CI: -1.61, -0.25) after adjusting for base model covariates. Similarly, at age 15 prenatal maternal smoking was associated with 0.93 percent lower DNA methylation at cg055975921 (95%CI: (-1.73, -0.13)). Prenatal maternal smoking remained significantly associated with a decrease in cg05575921 DNA methylation in the African genetic ancestry sample at age 9 (-1.33 (95%CI: -2.21, - 0.46)) and age 15 (-1.1 (95%CI: -2.14, -0.06); Figure 3). In the European genetic ancestry sample, the association was consistent in direction at age 9 (-0.85 (95%CI: - 2.47, 0.76)) and age 15 (-1.98 (95%CI: -3.7, -0.26)). In the Admixed-Latin genetic ancestry sample, the association was no longer significant and not consistent in direction (Age 9: 0.01 (95%CI: -1.67, 1.7); Age 15: 0.92 (95%CI: -1.19, 3.03)) (Figure 3).

Prenatal maternal smoking was not associated with global DNA methylation or epigenetic age (Figure 3).

### Sensitivity analysis: Specificity of DNA methylation signal to type and time of exposure

To assess the specificity of the DNA methylation signal to prenatal maternal cigarette smoking, we additionally adjusted for any other prenatal maternal drug use and prenatal maternal alcohol use. The association between prenatal maternal smoking and the polymethylation score was robust to this adjustment. The association between prenatal maternal smoking and cg05575921 was slightly attenuated (Supplemental Figure 4, Supplemental Table 3).

To assess the specificity of the DNA methylation signal to the timing of cigarette smoke exposure, we additionally adjusted for exposure to postnatal maternal smoking. Very few children exposed to maternal smoking prenatally were unexposed postnatally (Supplemental Figure 5). In bivariate analyses, children exposed to only postnatal smoking had lower polymethylation scores than those exposed to pre-and postnatal smoking. However, they did not have higher polymethylation scores than children unexposed to both pre-and postnatal smoking (Supplemental Figure 5). In multivariable analyses, prenatal maternal smoking was still associated with the polymethylation scores when adjusting for postnatal smoke exposure, though effect estimates were attenuated (Age 9: 0.39 (95%CI: 0.19, 0.59), Age 15: 0.33 (95%CI: 0.13, 0.54)). Prenatal maternal smoking was no longer associated with cg05575921 methylation after adjusting for postnatal smoke exposure (Age 9: -0.65 (95%CI: -1.52, 0.23), Age 15: - 0.37 (95%CI: -1.41, 0.67)).

Effect estimates were also similar when controlling for surrogate variables instead of known covariates, which corrects for both known and unknown sources of technical variation (Supplemental Figures 3 and 6; Supplemental Table 4). Effect estimates were also similar when additionally controlling for array position on DNA methylation slide in multivariable models with known covariates (data not shown).

### Sensitivity analysis: Choice of coefficients for constructing polymethylation score and associations with other a priori DNA methylation sites

Results were also similar when using sensitivity polymethylation scores (see Methods, Supplemental Figure 7).

In addition to cg055975921 in the *AHHR* gene, we tested the association of four other *a priori* DNA methylation sites identified from previous meta-analyses. We replicated the association of prenatal maternal smoking with DNA methylation at *GFI1*: cg14179389, *CYP1A1*: cg05549655, *MYO1G*: cg22132788, and *MYO1G*: cg04180046 (Supplemental Figure 8).

### Polymethylation scores improve classification of prenatal smoke exposure while AHRR: cg05575921 alone does not

Next, we assessed whether including DNA methylation summary measures increased the accuracy of classifying prenatal smoke exposure over a base model consisting of child sex, maternal income to poverty ratio at baseline, child age at DNA methylation measurement, estimated immune cell proportion, plate from methylation processing, the first two genetic principal components and an indicator variable for oversampled city (Figure 4). At age 9, including the polymethylation score increased the AUC by 5 (95% CI: (2.1, 7.2))) percentage points over the nested base model (*P*-value from DeLong test=0.0004). At age 15 including the polymethylation score for prenatal smoke exposure increased the AUC by 5 (95% CI: (2.1, 7.2)) percentage points (*P*-value from DeLong test=0.0004). Including DNA methylation at *AHRR*: cg0557591 with base model covariates did not improve the classification compared to using base model covariates alone (Supplemental Tables 5 and 6). Classification accuracy when including the polymethylation score from both age 9 and 15 was very similar to classification accuracy when using the polymethylation score from age 9 only (Figure 4B).

**Figure 4.**
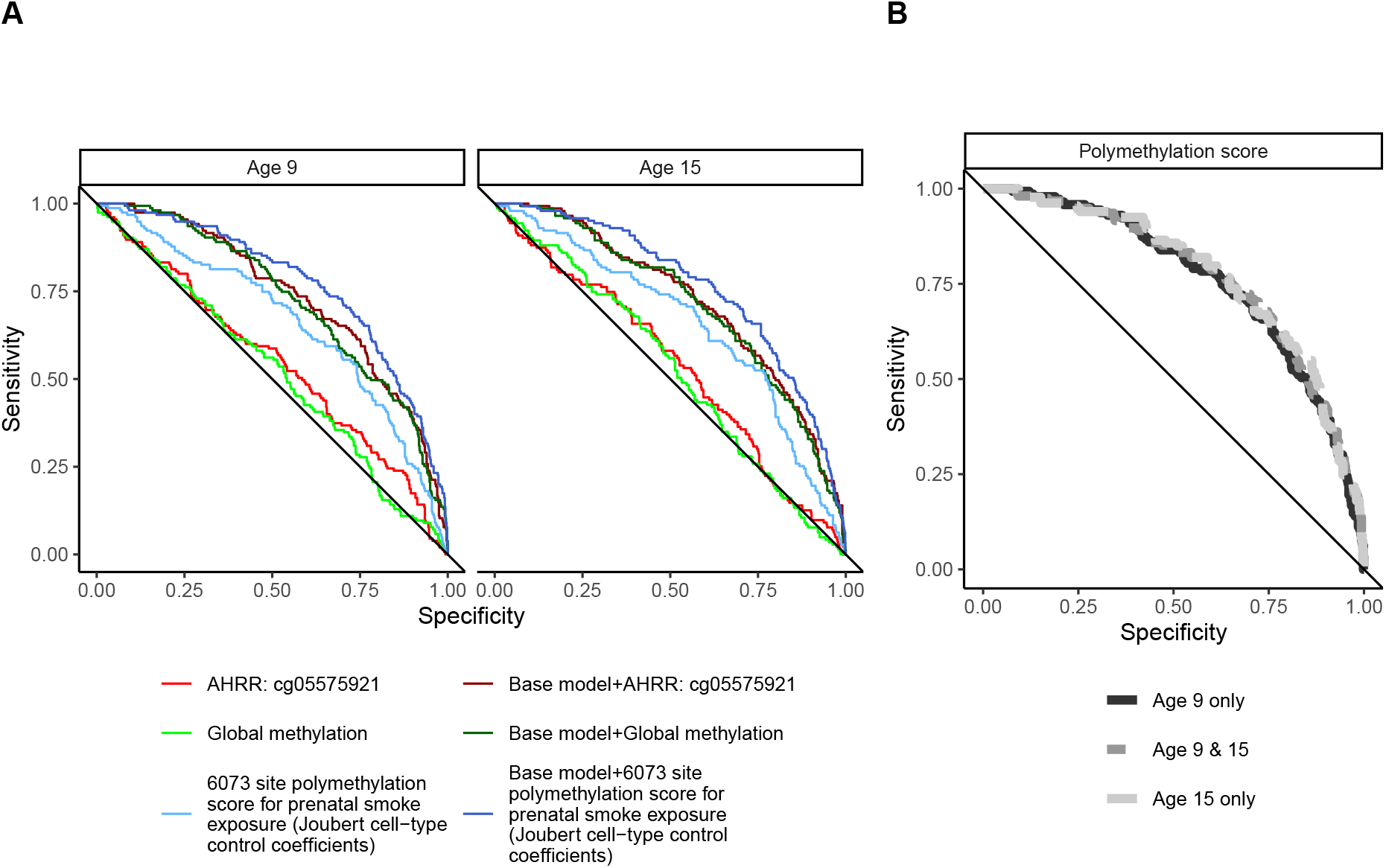
Polymethylation scores accurately classify prenatal maternal smoke exposure at ages 9 and 15 among 805 children in the Fragile Families and Child Wellbeing study. A) Receiver operator curve for select DNA methylation measures for predicting prenatal smoke exposure using no other variables (light colors) or using base model variables (dark colors, other variables included: child sex, maternal income-poverty ratio at birth, immune cell proportion and batch of methylation data processing). B) Receiver operator curve for include polymethylation scores individually at each visit (black & light grey) or jointly (dark grey).

### Sensitivity analysis: Impact of choice of coefficients on polymethylation score accuracy and comparison to AUCs reported in the literature

The AUC for classification of prenatal maternal smoke exposure in the Fragile Families study was similar across all polymethylation scores (Figure 5, Supplemental Table 6). The highest observed AUC was from the 568-site score (using strict coefficients from the regression of prenatal smoke exposure and DNA methylation in newborn cord blood). Performance was similar to previous reports. Neither the 28-site score from a LASSO regression nor the 204-site score from an elastic net regression performed as well in Fragile Families as has been reported previously in the literature (Figure 5).

**Figure 5.**
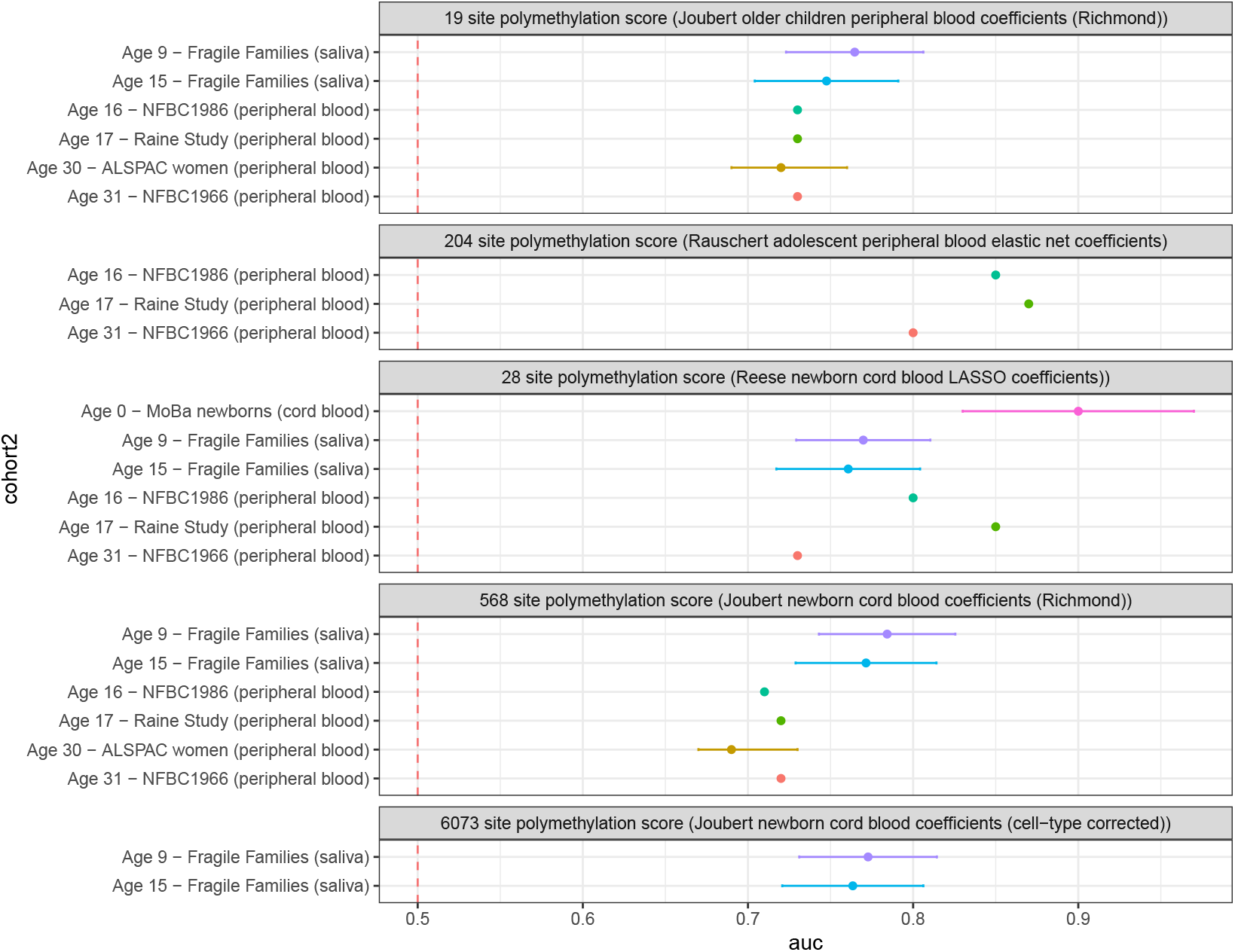
Comparison of different polymethylation scores and their accuracy at classifying prenatal smoke exposure among 805 children with saliva samples in the Fragile Families and Child Wellbeing study as opposed to previously reported results in cord and peripheral blood samples.

## Discussion

In the longitudinal Fragile Families and Child Wellbeing birth cohort, prenatal maternal smoking was associated with saliva DNA methylation in children at ages 9 and 15. Children with prenatal smoke exposure had 0.51 (95%CI: 0.35, 0.66) standard deviation higher polymethylation scores at age 9 than children not exposed. These findings were robust across strata of genetic ancestry and children’s age. These signatures were specific, as global DNA methylation and epigenetic clocks were not associated with prenatal smoke exposure. When assessing biomarker utility, the polymethylation score for prenatal smoke exposure improved the classification of self-reported prenatal maternal smoking over a base model by 5 (95% CI: (2.1, 7.2)) percentage points at age 9. A single *a priori* DNA methylation site, *AHRR*: cg05575921, did not (1 (95% CI: (-0.5, 2.6) percentage point increase). Our results suggest salivary DNA methylation can be used as an alternative to peripheral blood DNA methylation for biomarkers of prenatal maternal smoking. A saliva-based DNA methylation biomarker may be particularly useful in cases where blood collection, processing, and storage is not possible. Saliva can often be collected directly by participants and saliva sample collection kits have stabilizers allowing room temperature storage prior to DNA methylation measurement.

Our findings are consistent with the previous literature in different tissues and at different ages on associations between prenatal maternal smoking and DNA methylation. We replicated top hits from the seminal epigenome wide association meta-analysis of prenatal smoke exposure and DNA methylation of cord blood from newborns including *AHRR*: cg05575921, *GFl1*: cg14179389 and *MYO1G1*: cg22132788. Consistent with a meta-analysis of peripheral blood from children and adults over 16 years of age, the association at *MYO1G1*: cg22132788 was larger than in newborn cord blood [9, 35]. Again, consistent with a meta-analysis of peripheral blood from children and adults over 16 years of age, the associations at *AHRR*: cg05575921 and *GFl1*: cg14179389 were smaller than in newborn cord blood [9, 35]. There are no previous studies of prenatal maternal smoking and saliva DNA methylation. However, immune cell types are present in both blood and saliva, and saliva samples in our study were primarily composed of immune cells. Additionally, fewer than 4% of DNA methylation sites in blood and saliva samples displayed significantly different methylation values (|*Δβ*| > 0.2 and *p*_*adj*_ < 0.001) when measured on the 450K chip in samples of adults [41] and 11-year-old children [14].

We also advance the prenatal smoking - DNA methylation literature by evaluating the generalizability of association between prenatal maternal smoking and DNA methylation across genetic ancestry groups. The portability of polygenic risk scores across genetic ancestries is a complex research area [42] and evaluating the portability of epigenetic summary measures has been identified as a key area for evaluation [43]. Our results suggest polymethylation scores for prenatal maternal smoking are portable across genetic ancestry groups. At age 9, the effect estimates for prenatal maternal smoking on the polymethylation score in the European and African genetic ancestry samples were within 0.01 standard deviation and were both significant at an alpha of 0.05. This finding is in line with a recent epigenome-wide association study in which children of both black and non-black identifying mothers exhibited similar directions of associations between prenatal smoke exposure and peripheral blood methylation at 38 DNA methylation sites [44]. In our analysis, the association between prenatal smoke exposure and polymethylation scores was attenuated in children of admixed genetic ancestry (primarily of Hispanic ethnicity) at both ages and in children of European genetic ancestry at age 15. This could reflect differences in DNA methylation by genetic sequence at prenatal smoke sensitive DNA methylation sites [18]. While genetic ancestry is distinct from racial and ethnic social and cultural constructs [45], it can correlate with different social and environmental exposures. Differences in other social and environmental exposures could also contribute to the attenuation observed in the admixed genetic ancestry group.

We contribute to the understanding of the impact of tissue on classification accuracy of DNA methylation biomarkers for prenatal maternal smoking. Polymethylation scores using coefficients from the Joubert meta-analysis coefficients had similar AUCs in Fragile Families saliva samples (AUCs between 0.61 and 0.78) and in previous applications in peripheral blood samples from ages 17-, 30- and 31-years (AUCs between 0.69 and 0.72) [12, 33]. Thus, using polymethylation scores from saliva may be an acceptable alternative when peripheral blood samples are unavailable or difficult to obtain. However, other polymethylation scores for prenatal maternal smoking have performed better in peripheral blood samples. The greatest classification accuracy of prenatal smoke exposure from previous studies was with a 28-site LASSO regression score trained in and then applied to a Norwegian cord blood cohort (AUC=0.90) [21]. This score also accurately classified prenatal smoking from peripheral blood from adolescents and adults (AUCs between 0.85 and 0.73) [33]. A 204-site elastic net score trained and then applied to peripheral blood from 17-year-old’s peripheral blood samples also performed very well (AUC=0.87) [33]. This score also performed well when applied to additional validation cohorts of peripheral blood from adolescents and adults (AUCs between 0.8 and 0.87). However, neither of these scores performed as well when applied in the Fragile Families cohort. The difference in classification accuracy could reflect cell proportion differences across tissues. Using coefficients from an epigenome-wide association analysis of saliva could improve classification and is a direction for future research.

While polymethylation scores significantly improved classification of prenatal smoke exposure over a base model, using *AHRR*: cg05575921 methylation as a classifier did not (percentage point increase in AUC: 1 (95% CI: (-0.5, 2.6)))). Prenatal smoke exposed newborns consistently exhibit lower DNA methylation at *AHRR*: cg05575291 than unexposed newborns [9]. However, the association between prenatal smoke exposure and *AHRR*: cg05575291 methylation did not persist in peripheral blood samples from adult women [12]. Additionally, a previous longitudinal analysis of children’s blood samples from birth to 17 years of age in the ALSPAC cohort found that the prenatal smoke exposure difference in DNA methylation at AHRR: cg05575291 had the highest magnitude at birth, with attenuated differences at 17 years of age [12]. The attenuated differences in DNA methylation at AHRR: cg05575291 in adolescence was partially mediated by postnatal own-smoking [12], thus in our analysis we excluded the few children who reported postnatal own-smoking. They observed that prenatal smoke exposure associated DNA methylation differences at other sites were more persistent throughout development [12]. As the persistence and accuracy of *AHRR*: cg05575921 and other single CpG site biomarkers may be influenced by time-since-exposure and new environmental exposures, incorporating information across multiple sites of DNA methylation may yield a more robust biomarker.

Additionally, we found that prenatal maternal smoke exposure was not associated with global DNA methylation or a pediatric saliva and the GrimAge DNA methylation clocks, underlining the specificity of the DNA methylation signature of prenatal smoke exposure. DNA methylation clocks attempt to measure aspects of biological aging and are trained in specific populations. We note that the pediatric DNA methylation clock, which was specifically trained for use in pediatric populations, accurately recapitulated our subjects chronological ages. However, the GRIM age clock, which was trained in adults to measure proximity to mortality, overestimated chronological ages in our population by 10-15 years. In an analysis of four pediatric cohorts of children under 20 years of age, GRIM age estimates were 30-35 years greater than chronological in buccal cell samples and 10-15 years greater than chronological age in blood samples [46]. GRIM age was neither trained nor validated in a pediatric population [37], which may explain this overestimation.

Our analysis is not without its limitations. We used maternal self-report of prenatal maternal smoke exposure, as serum cotinine levels were not available. As mothers may be reluctant to report smoking during pregnancy, some of our children may be misclassified as unexposed to prenatal smoke. We would expect this to bias our results towards the null. Analyses of prenatal exposures and postnatal outcomes are conditioned on live birth and survival or enrollment until the point of outcome measurement. Because prenatal maternal smoking is associated with miscarriage [47], this could result in selection bias if an unmeasured variable is associated with both live birth and the outcome. This selection bias can create a downward bias in the effect estimate. This could produce a spurious protective effect (which we did not observe), or, in the case of a true positive association between exposure and outcome, will attenuate the association toward the null [48, 49]. Additionally, while we controlled for postnatal secondhand smoke exposure and excluded children who reported any own-smoking, we cannot exclude the possibility some of the observed effects are mediated through these alternative sources of smoke exposure. Personal smoking behavior among children in our cohort was relatively rare, and we excluded those participants from our analysis to focus on the prenatal exposure window. Future studies may focus on enriched risk samples to examine the joint effect of prenatal smoke exposure and personal smoking. In adults, DNA methylation has also been associated with BMI, alcohol consumption, and inflammation [50–53]. Future work could also investigate how these postnatal exposures associate with prenatal smoking and DNA methylation in a joint exposure or mixtures analysis. While we demonstrate that changes to DNA methylation observed in children’s cord blood can still be detected in saliva samples from adolescence, future studies of adults will be necessary to confirm if these changes persist to adulthood.

Our analysis is strengthened by our prospective study design, diverse study participants sampled, rigorous outcome measures, and advanced analytics. We analyzed samples from a large cohort of diverse participants, with African, Admixed-Latin, and European genetic ancestry participants, including those who are currently underrepresented in genetic and epigenetic research [54, 55]. Our measurement of prenatal maternal smoke exposure was prospectively assessed at birth and preceded outcome measurements of DNA methylation measures, limiting the potential for recall bias. We analyzed repeated measures of DNA methylation with reproducible array measures conducted in a single batch, reducing the impacts of batch effects. We tested associations between prenatal maternal smoking and multiple DNA summary measures to evaluate the specificity of the polymethylation scores. To examine the specificity of the biomarker to the nature and timing of exposure, we adjusted for prenatal maternal drinking, other prenatal maternal drug use, and postnatal secondhand smoke exposure.

## Conclusions

In a large, prospective study of diverse participants, we showed that DNA methylation in children’s saliva had strong associations with and reasonable classification accuracy for prenatal maternal smoke exposure. Further, we demonstrated that polymethylation scores could be applied as a biomarker of prenatal maternal smoke exposure across genetic ancestry groups, an important consideration for the equitable biomarker development. The development and application of biomarkers for prenatal maternal smoke exposure has important implications for epidemiological research and clinical practice. Given the difficulty of measuring prenatal maternal smoke exposure, such a biomarker could allow for confounder control in research areas where such control is currently impossible. Prenatal maternal smoke exposure is prevalent and has negative health consequences for developing fetuses, including associations with postnatal conditions such as obesity, asthma, and neurodevelopmental disorders [1–3]. Thus, a salivary exposure biomarker could be used to identify exposed children at risk of these outcomes and provide support and health interventions.

## Data Availability

Code to perform all analyses is available (www.github.com/bakulskilab). Survey data for the Fragile Families and Child Wellbeing study are publicly available (https://fragilefamilies.princeton.edu/documentation). DNA methylation and genetic data in the Fragile Families and Child Wellbeing study are available through the Restricted Use Contract (https://fragilefamilies.princeton.edu/restricted), upon demonstrated Institutional Review Board approval of the proposed research and data protection plan.

## Declarations

### Ethics approval and consent to participate

Participants provided written informed consent for the study. The data used in this manuscript were prepared by the Fragile Families and Childhood Wellbeing Study administrators following approval of the manuscript proposal. These secondary data analyses were approved by the University of Michigan Institutional Review Board (IRB, HUM00129826).

### Consent for publication

Not applicable

### Authors’ contributions

FB performed the analysis with contributions from JD, JF, and EBW. FB and KB wrote the paper with all authors contributing to revisions. CM, DN and LS contributed to the design and processing of the DNA methylation data subcohort. DN, CM, KM, EBW, and FB all contributed to the design and conception of the analytic plan. All authors reviewed and revised the manuscript.

## Acknowledgements

Not applicable

