## Supplemental Tables 2-6 and Supplemental Figures 1-8 for "Polymethylation Scores for Prenatal Maternal Smoke Exposure Persist Until Age 15 and Are Detected in Saliva in the Fragile Families and Child Wellbeing Cohort"

*Senior co-authors

^+^Corresponding author

Supplemental Table 2 - Descriptive statistics of excluded vs included samples with methylation data available from the Fragile and Families and Child Wellbeing study. Samples excluded according to the flow diagram in Main Figure 1

| **Characteristic** | M samples | **Final analysis group**,  M samples = 1500*^1^* | **Not in final analysis group**,  M samples = 184*^1^* |
| --- | --- | --- | --- |
| Prenatal maternal smoking | 1,684 |  |  |
| Not exposed |  | 1,202 (80%) | 141 (77%) |
| Smoke exposed |  | 298 (20%) | 43 (23%) |
| Age at DNA methylation sample | 1,684 |  |  |
| Age 15 |  | 747 (50%) | 109 (59%) |
| Age 9 |  | 753 (50%) | 75 (41%) |
| 6073 site polymethylation score: coefficients for sustained smoking from Joubert newborn cord blood meta-analysis, w/ cell-type control - PMC4833289 (mean-centered) | 1,684 | 0.00 (0.25) | -0.01 (0.27) |
| 568 site polymethylation score (Richmond): coefficients for sustained smoking from Joubert newborn cord blood meta-analysis - PMC4833289 (mean-centered) | 1,684 | 0.00 (0.08) | 0.00 (0.08) |
| 28 site polymethylation score (Reese): coefficients for sustained smoking from newborn cord blood LASSO regression - PMC5381987 (mean-centered) | 1,684 | -0.04 (2.31) | 0.31 (2.29) |
| 204 site polymethylation score classification probability (Rauschert): coefficients for sustained smoking from older children peripheral blood elastic net regression | 1,684 | 0.52 (0.14) | 0.52 (0.15) |
| *AHRR*: cg05575921 | 1,684 | 76.9 (5.9) | 74.9 (7.5) |
| Pediatric clock | 1,684 | 11.51 (2.66) | 11.83 (2.55) |
| Global methylation | 1,684 | 50.27 (1.02) | 50.12 (1.24) |
| Immune cell proportion (saliva) | 1,684 | 0.94 (0.13) | 0.93 (0.15) |
| Epithelial cell proportion (saliva) | 1,684 | 0.06 (0.13) | 0.07 (0.15) |
| Ancestry categorization from child principal components of genetic data | 1,665 |  |  |
| Admixed ancestry - Latin heritage |  | 351 (23%) | 68 (41%) |
| African ancestry |  | 911 (61%) | 78 (47%) |
| European ancestry |  | 238 (16%) | 19 (12%) |
| Unknown |  | 0 | 19 |
| Child gender | 1,684 |  |  |
| Boy |  | 741 (49%) | 106 (58%) |
| Girl |  | 759 (51%) | 78 (42%) |
| Oversampled cities | 1,684 |  |  |
| Detroit, Chicago or Toledo |  | 417 (28%) | 55 (30%) |
| Not Detroit, Chicago or Toledo |  | 1,083 (72%) | 129 (70%) |
| Child own smoking asked at age 15: Ever smoked an entire cigarette? | 1,666 |  |  |
| Yes |  | 32 (2.1%) | 49 (28%) |
| No |  | 1,460 (98%) | 125 (72%) |
| Unknown |  | 8 | 10 |
| Child own smoking asked at age 9: Smoked a cigarette or used tobacco | 1,662 |  |  |
| Yes |  | 0 (0%) | 12 (6.8%) |
| No |  | 1,485 (100%) | 165 (93%) |
| Unknown |  | 15 | 7 |
| Maternal prenatal alcohol use | 1,680 | 174 (12%) | 20 (11%) |
| Unknown |  | 0 | 4 |
| Maternal prenatal any drug use | 1,678 | 73 (4.9%) | 12 (6.7%) |
| Unknown |  | 0 | 6 |
| Constructed - Poverty ratio - mother's household income/poverty threshold | 1,684 | 2.25 (2.46) | 2.37 (2.56) |
| Any postnatal maternal smoking when child age 1 or 5 | 1,596 |  |  |
| Maternal smoking at age 1 or age 5 |  | 588 (39%) | 50 (52%) |
| No maternal smoking at age 1 and 5 |  | 912 (61%) | 46 (48%) |
| Unknown |  | 0 | 88 |
| Maternal/primary care giver smoking in month prior to visit (pks/day) | 1,666 |  |  |
| Less than pack a day |  | 339 (23%) | 42 (25%) |
| No smoking |  | 1,052 (70%) | 115 (69%) |
| Pack or more a day |  | 109 (7.3%) | 9 (5.4%) |
| Unknown |  | 0 | 18 |
| *^1^* n (%); Mean (SD) | | | |

Supplemental Figure 1 - Pearson correlation of selected DNA methylation summary measures from an analysis of M=1500 samples and N=805 children in the Fragile Families and Child Wellbeing cohort. Lower diagonal shows the Pearson rho correlation coefficient, upper triangle shows ellipses illustrating relationship between variables colored by the magnitude and direction of the correlation. Immune cell and epithelial cell proportions were calculated from DNA methylation measures in children’s saliva and a children’s saliva reference panel.

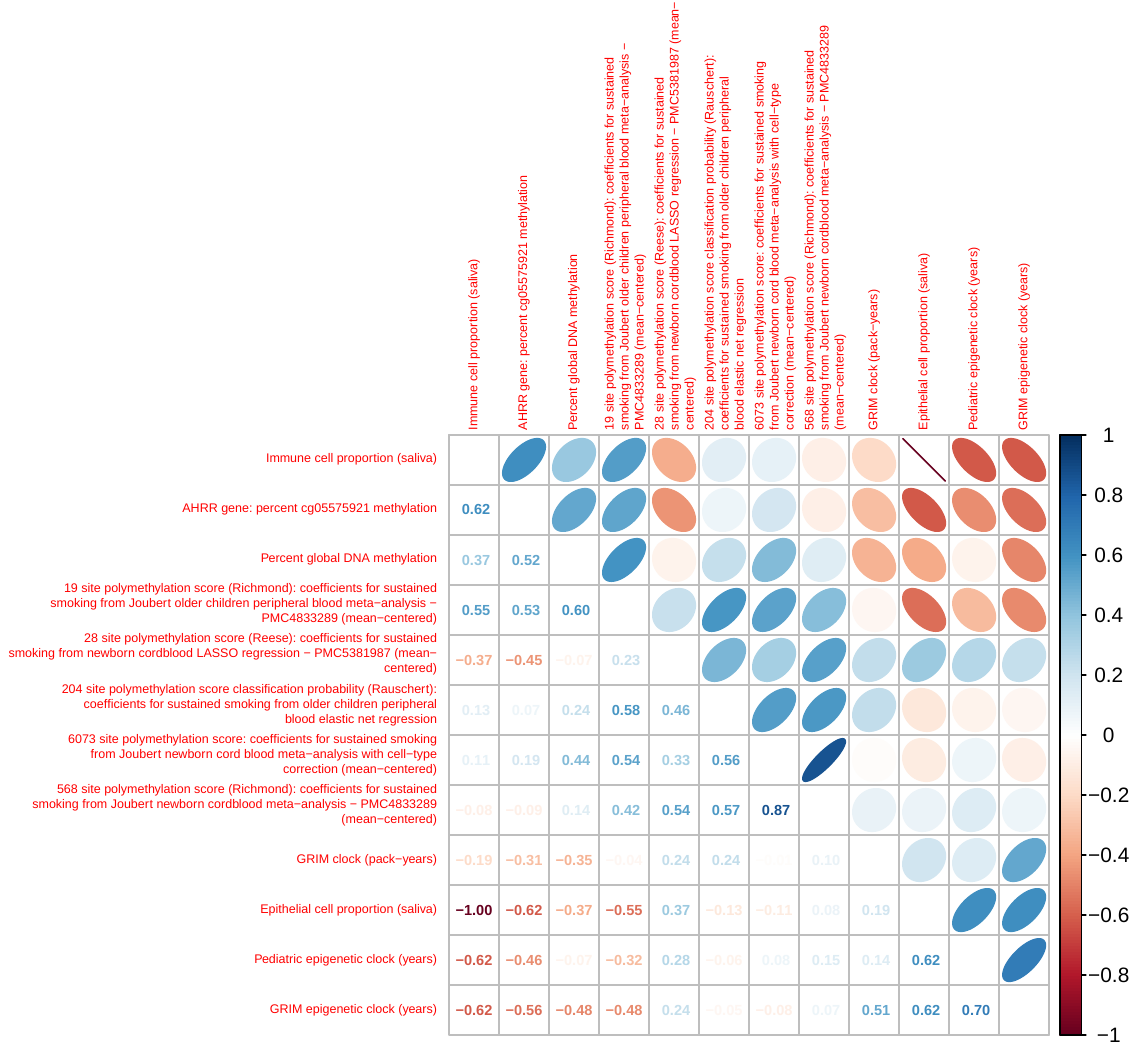

Supplemental Figure 2 - Correlation between age 9 and age 15 visit DNA methylation summary measures among 695 children with age 9 and age 15 visit data in the Fragile Families and Child Wellbeing study. Each point represents one individual, x-axis is the DNA methylation summary measure at age 9 and y-axis is the DNA methylation measure at age 15. Blue lines are simple linear regression lines of best fit, with Pearson rho and associated *P* value shown. From top-left: Polymethylation score for prenatal maternal smoking, score created using coefficients from Joubert meta-analysis regression of prenatal maternal smoking and DNA methylation in newborn cord blood, incorporating cell-type control (6073 DNA methylation sites). Polymethylation score for prenatal maternal smoking using coefficients Joubert meta-analysis regression of prenatal maternal smoking and DNA methylation in newborn cord blood, no cell-type control (568 DNA methylation sites). Polymethylation score calculated using Joubert meta-analysis regression coefficients from a regression of prenatal maternal smoking and DNA methylation in older children’s peripheral blood samples (19 DNA methylation sites). Polymethylation score calculated using coefficients from a LASSO regression of prenatal maternal smoking and DNA methylation in children’s cord blood (28 DNA methylation sites). Polymethylation score (classification probability) calculated using elastic net regression from adolescent peripheral blood samples (204 DNA methylation sites). Pediatric epigenetic clock (years). GRIM epigenetic clock (years). Percent DNA methylation at cg05575921 in AHRR. Percent global DNA methylation (mean across all DNA methylation sites in cleaned probe set). Proportion of epithelial cells calculated from DNA methylation using reference panel of children’s saliva. Proportion of immune cells calculated from DNA methylation using a reference panel of children’s saliva.

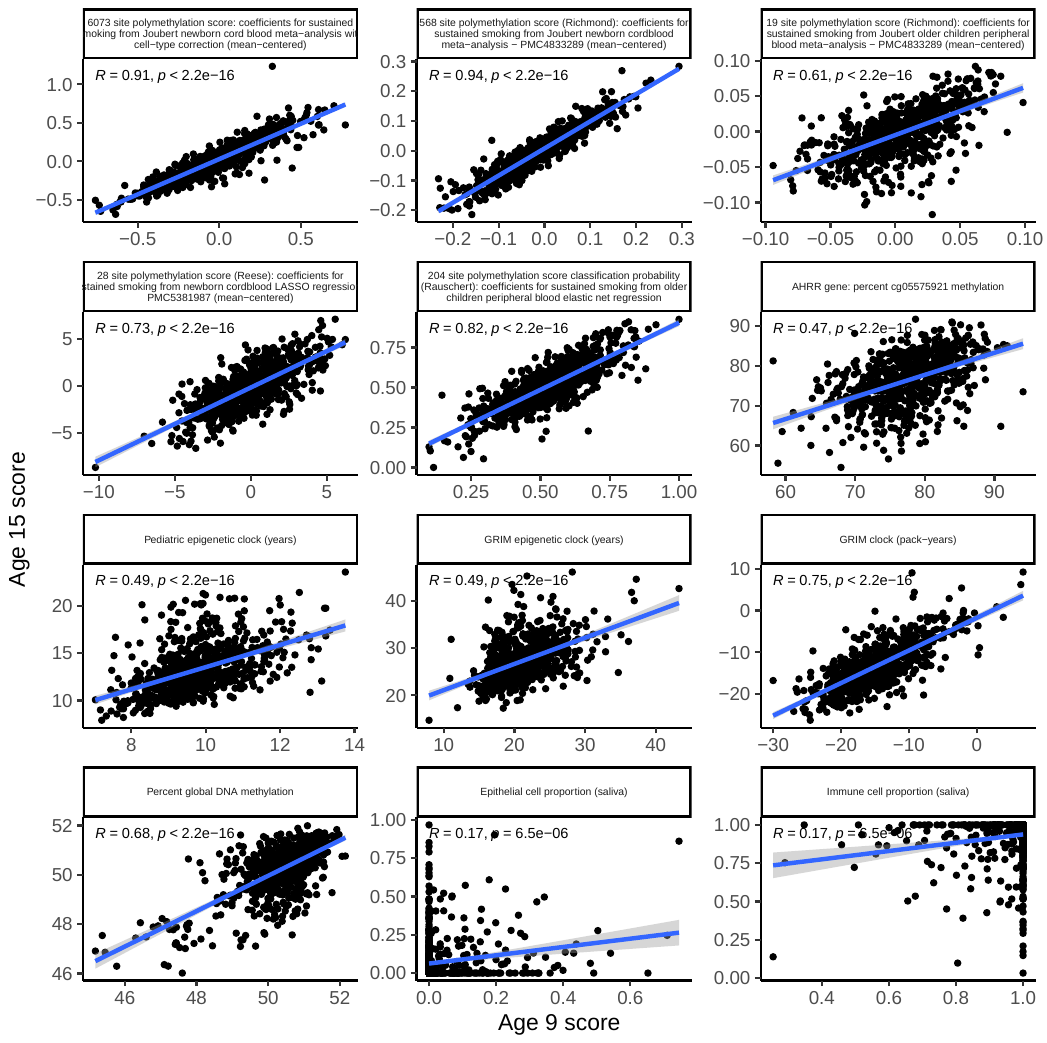
Supplemental Figure 3: Distributions of selected methylation summary measures at age 9 and age 15 among 805 children in the Fragile Families and Child Wellbeing study. Captive box plots within violin plots show the distribution of selected DNA methylation measures in samples from age 9 (red) and age 15 (blue). Black lines show lines of best fit. From top-left: Polymethylation score for prenatal maternal smoking, score created using coefficients from Joubert meta-analysis regression of prenatal maternal smoking and DNA methylation in newborn cord blood, incorporating cell-type control (6073 DNA methylation sites). Polymethylation score for prenatal maternal smoking using coefficients Joubert meta-analysis regression of prenatal maternal smoking and DNA methylation in newborn cord blood, no cell-type control (568 DNA methylation sites). Polymethylation score calculated using Joubert meta-analysis regression coefficients from a regression of prenatal maternal smoking and DNA methylation in older children’s peripheral blood samples (19 DNA methylation sites). Polymethylation score calculated using coefficients from a LASSO regression of prenatal maternal smoking and DNA methylation in children’s cord blood (28 DNA methylation sites). Polymethylation score (classification probability) calculated using elastic net regression from adolescent peripheral blood samples (204 DNA methylation sites). Pediatric epigenetic clock (years). GRIM epigenetic clock (years). Percent DNA methylation at cg05575921 in AHRR. Percent global DNA methylation (mean across all DNA methylation sites in cleaned probe set). Proportion of epithelial cells calculated from DNA methylation using reference panel of children’s saliva. Proportion of immune cells calculated from DNA methylation using a reference panel of children’s saliva.
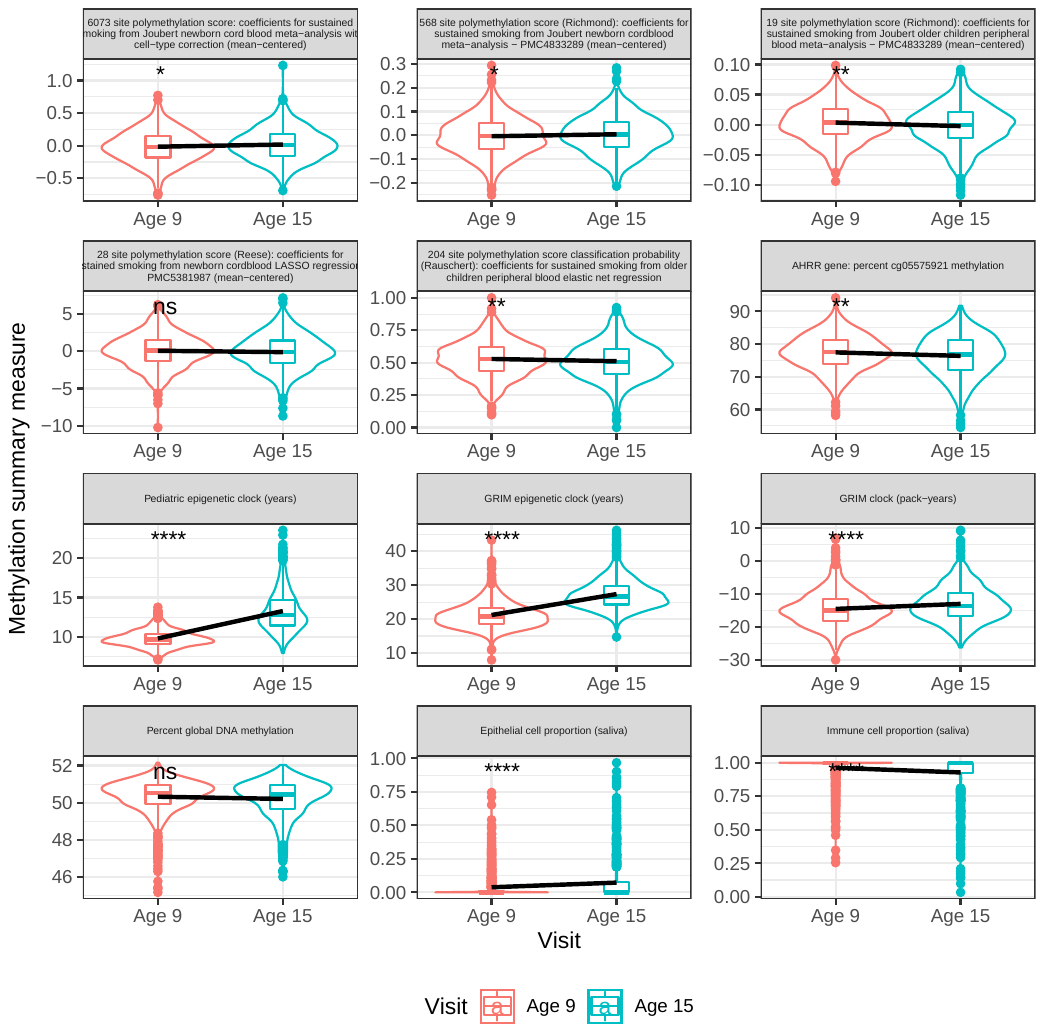

Supplemental Table 3: Bivariate associations between prenatal maternal smoking and additional DNA methylation summary measures among a diverse sample of 805 children in the Fragile Families and Child Wellbeing study. Percent DNA methylation shown calculated across all DNA methylation sites available (global DNA methylation) and stratified by genomic region (Island, Open Sea, Shelf, Shore). Polymethylation score for smoke exposure from main analysis and from sensitivity analyses where scores were calculated using alternative available regression coefficients.

| **Characteristic** | **Age 9** | | | | **Age 15** | | | |
| --- | --- | --- | --- | --- | --- | --- | --- | --- |
|  | **N** | **Not exposed**, N = 598*^1^* | **Smoke exposed**, N = 155*^1^* | **p-value***^2^* | **N** | **Not exposed**, N = 604*^1^* | **Smoke exposed**, N = 143*^1^* | **p-value***^2^* |
| Percent global DNA methylation (%) | 753 | 50.32 (0.95) | 50.34 (1.08) | 0.9 | 747 | 50.19 (1.07) | 50.28 (1.01) | 0.3 |
| Island methylation (%) | 753 | 19.44 (0.61) | 19.49 (0.61) | 0.4 | 747 | 19.53 (0.61) | 19.57 (0.58) | 0.4 |
| Open Sea methylation (%) | 753 | 74.99 (1.41) | 74.97 (1.60) | >0.9 | 747 | 74.69 (1.61) | 74.82 (1.50) | 0.3 |
| Shelf methylation (%) | 753 | 80.06 (1.45) | 80.02 (1.65) | 0.8 | 747 | 79.75 (1.64) | 79.87 (1.52) | 0.4 |
| Shore methylation (%) | 753 | 46.88 (1.01) | 46.92 (1.16) | 0.7 | 747 | 46.75 (1.15) | 46.84 (1.09) | 0.4 |
| Pediatric epigenetic clock (years) | 753 | 9.80 (1.07) | 9.70 (1.13) | 0.3 | 747 | 13.27 (2.64) | 13.24 (2.57) | 0.9 |
| GRIM epigenetic clock (years) | 753 | 21.2 (4.0) | 21.0 (3.8) | 0.4 | 747 | 27.4 (4.5) | 27.1 (4.5) | 0.5 |
| GRIM clock smoking subcomponent (pack-years) | 753 | -14.8 (5.2) | -13.8 (5.2) | 0.039 | 747 | -13.1 (5.5) | -12.8 (5.3) | 0.5 |
| Epithelial cell proportion (saliva) | 753 | 0.04 (0.10) | 0.03 (0.08) | 0.3 | 747 | 0.07 (0.15) | 0.07 (0.14) | 0.9 |
| Immune cell proportion (saliva) | 753 | 0.96 (0.10) | 0.97 (0.08) | 0.3 | 747 | 0.93 (0.15) | 0.93 (0.14) | 0.9 |
| Polymethylation score for smoke exposure (primary outcome): 6073 site score using coefficients from Joubert newborn cord blood meta-analysis with cell-type control | 753 | -0.04 (0.24) | 0.10 (0.25) | <0.001 | 747 | -0.01 (0.25) | 0.13 (0.25) | <0.001 |
| 568 site polymethylation score (Richmond): coefficients for sustained smoking from Joubert newborn cord blood meta-analysis - PMC4833289 (mean-centered) | 753 | -0.02 (0.08) | 0.04 (0.08) | <0.001 | 747 | -0.01 (0.08) | 0.05 (0.08) | <0.001 |
| 19 site polymethylation score (Richmond): coefficients for sustained smoking from Joubert older children peripheral blood meta-analysis - PMC4833289 (mean-centered) | 753 | 0.00 (0.03) | 0.02 (0.03) | <0.001 | 747 | 0.00 (0.03) | 0.01 (0.04) | <0.001 |
| 28 site polymethylation score (Reese): coefficients for sustained smoking from newborn cord blood LASSO regression - PMC5381987 (mean-centered) | 753 | -0.20 (2.19) | 1.03 (2.20) | <0.001 | 747 | -0.36 (2.32) | 0.86 (2.31) | <0.001 |
| 204 site polymethylation score classification probability (Rauschert): coefficients for sustained smoking from older children peripheral blood elastic net regression | 753 | 0.51 (0.13) | 0.60 (0.14) | <0.001 | 747 | 0.50 (0.14) | 0.58 (0.15) | <0.001 |
| *AHRR* gene: percent cg05575921 methylation | 753 | 77.6 (5.4) | 76.7 (5.6) | 0.070 | 747 | 77 (6) | 76 (6) | 0.2 |
| *MYO1G:* percent cg04180046 methylation | 753 | 42 (10) | 48 (12) | <0.001 | 747 | 41 (11) | 46 (12) | <0.001 |
| *CYP1A1*: percent cg05549655 methylation | 753 | 18 (7) | 21 (7) | <0.001 | 747 | 17 (7) | 20 (7) | <0.001 |
| *GFI1:* percent cg14179389 methylation | 753 | 4.80 (2.74) | 3.99 (2.23) | <0.001 | 747 | 4.91 (2.77) | 3.93 (1.70) | <0.001 |
| *MYO1G:* percent cg22132788 methylation | 753 | 69 (10) | 73 (9) | <0.001 | 747 | 67 (12) | 70 (12) | 0.003 |
| *^1^* Mean (SD)  *^2^* Welch Two Sample t-test | | | | | | | | |

Supplemental Figure 4: Sensitivity analyses for controlling for different variable sets in multivariable analyses of prenatal maternal smoking and DNA methylation summary measures. Base model: Child sex, maternal income to poverty ratio, immune cell proportion, plate, 1st two principal components from PCA on child genetic data, child age, indicator variable for over sampled cities. Prenatal exposures model: base model + yes/no any prenatal maternal alcohol use, yes/no any prenatal other drug use. Secondhand smoking models: prenatal exposures model + yes/no any postnatal maternal/primary care giver smoking at age 1 or 5, pk/day in month prior to primary caregiver/maternal interview at age 9 or 15. Surrogate variable: including all surrogate variables

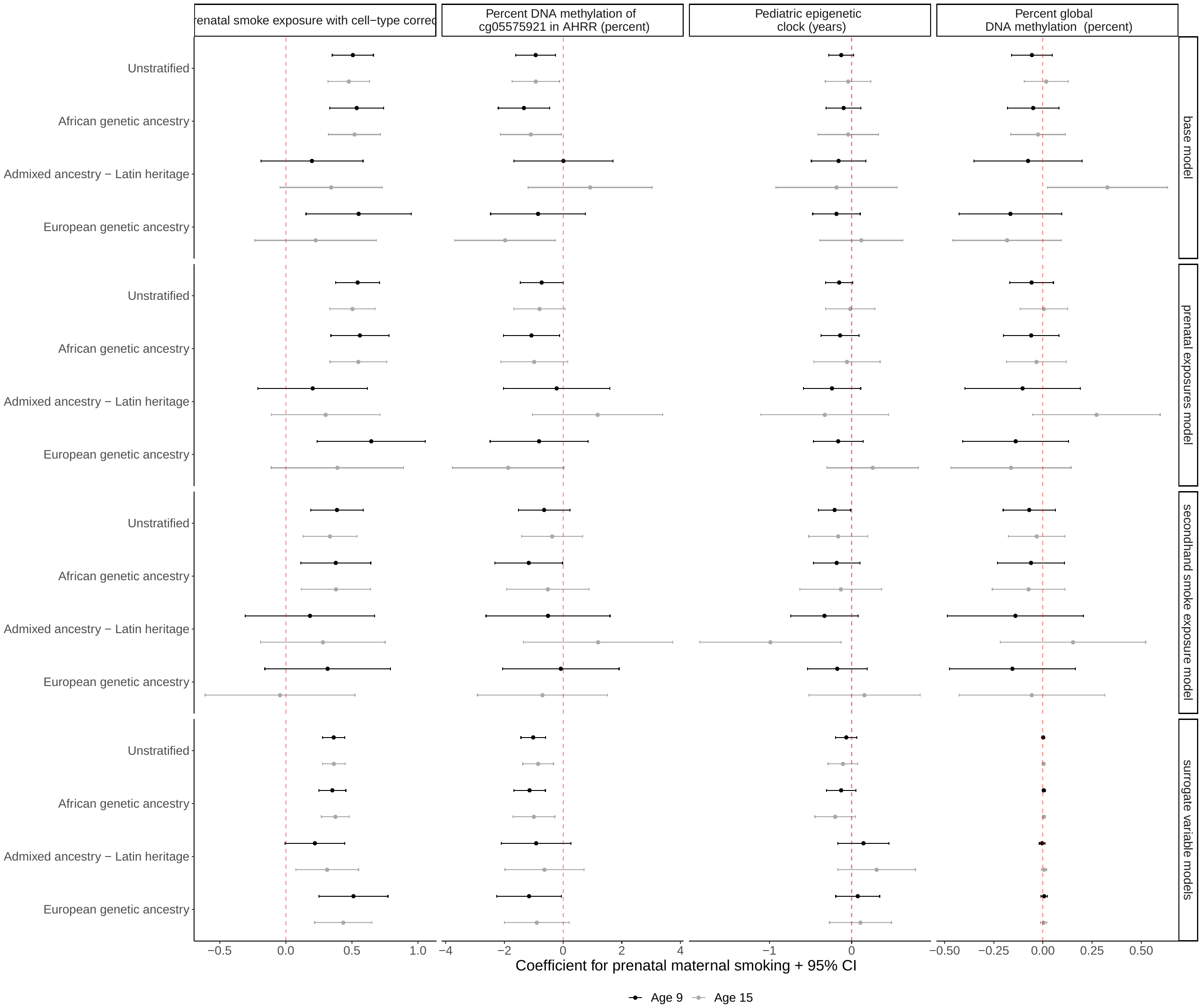

Supplemental Table 4: Associations between prenatal maternal smoking and DNA methylation summary measures in multivariable linear models stratified by age and/or ancestry and controlling for specified covariate sets. Base model: Child sex, maternal income to poverty ratio, immune cell proportion, plate, 1st two principal components from PCA on child genetic data, child age, indicator variable for over sampled cities. Prenatal exposures model: base model + yes/no any prenatal maternal alcohol use, yes/no any prenatal other drug use. Secondhand smoking models: prenatal exposures model + yes/no any postnatal maternal/primary care giver smoking at age 1 or 5, pk/day in month prior to primary caregiver/maternal interview at age 9 or 15. Surrogate variable: including all surrogate variables

| **DNA methylation summary measure (outcome variable)** | **Age strata** | **Genetic ancestry strata** | **Model type/covariate set** | **N** | **Estimate for prenatal maternal smoking** | **Lower 95% CI** | **Upper 95% CI** | **p.value** |
| --- | --- | --- | --- | --- | --- | --- | --- | --- |
| Global methylation | Age 15 | Admixed ancestry - Latin heritage | base model | 171 | 0.33 | 0.023 | 0.63 | 0.035383 |
| Global methylation | Age 15 | Admixed ancestry - Latin heritage | prenatal exposures model | 171 | 0.27 | -0.051 | 0.6 | 0.097667 |
| Global methylation | Age 15 | Admixed ancestry - Latin heritage | secondhand smoke exposure model | 171 | 0.15 | -0.22 | 0.52 | 0.41437 |
| Global methylation | Age 15 | Admixed ancestry - Latin heritage | surrogate variable models | 171 | 0.0063 | -0.0074 | 0.02 | 0.36575 |
| Global methylation | Age 15 | African ancestry | base model | 457 | -0.025 | -0.16 | 0.11 | 0.72652 |
| Global methylation | Age 15 | African ancestry | prenatal exposures model | 457 | -0.032 | -0.18 | 0.12 | 0.67439 |
| Global methylation | Age 15 | African ancestry | secondhand smoke exposure model | 457 | -0.073 | -0.26 | 0.11 | 0.43667 |
| Global methylation | Age 15 | African ancestry | surrogate variable models | 457 | 0.005 | -0.0027 | 0.013 | 0.20566 |
| Global methylation | Age 15 | European ancestry | base model | 119 | -0.18 | -0.46 | 0.093 | 0.19265 |
| Global methylation | Age 15 | European ancestry | prenatal exposures model | 119 | -0.16 | -0.47 | 0.14 | 0.29387 |
| Global methylation | Age 15 | European ancestry | secondhand smoke exposure model | 119 | -0.057 | -0.43 | 0.31 | 0.76206 |
| Global methylation | Age 15 | European ancestry | surrogate variable models | 119 | 0.0038 | -0.012 | 0.02 | 0.63591 |
| Global methylation | Age 15 | Multiethnic sample | base model | 747 | 0.017 | -0.094 | 0.13 | 0.76758 |
| Global methylation | Age 15 | Multiethnic sample | prenatal exposures model | 747 | 0.0045 | -0.12 | 0.13 | 0.94137 |
| Global methylation | Age 15 | Multiethnic sample | secondhand smoke exposure model | 747 | -0.031 | -0.17 | 0.11 | 0.66754 |
| Global methylation | Age 15 | Multiethnic sample | surrogate variable models | 747 | 0.0035 | -0.0025 | 0.0095 | 0.24849 |
| Global methylation | Age 9 | Admixed ancestry - Latin heritage | base model | 180 | -0.075 | -0.35 | 0.2 | 0.58923 |
| Global methylation | Age 9 | Admixed ancestry - Latin heritage | prenatal exposures model | 180 | -0.1 | -0.4 | 0.19 | 0.48852 |
| Global methylation | Age 9 | Admixed ancestry - Latin heritage | secondhand smoke exposure model | 180 | -0.14 | -0.49 | 0.21 | 0.42699 |
| Global methylation | Age 9 | Admixed ancestry - Latin heritage | surrogate variable models | 180 | -0.0042 | -0.018 | 0.01 | 0.56142 |
| Global methylation | Age 9 | African ancestry | base model | 454 | -0.05 | -0.18 | 0.081 | 0.45629 |
| Global methylation | Age 9 | African ancestry | prenatal exposures model | 454 | -0.06 | -0.2 | 0.082 | 0.40841 |
| Global methylation | Age 9 | African ancestry | secondhand smoke exposure model | 454 | -0.06 | -0.23 | 0.11 | 0.48735 |
| Global methylation | Age 9 | African ancestry | surrogate variable models | 454 | 0.004 | -0.0039 | 0.012 | 0.31793 |
| Global methylation | Age 9 | European ancestry | base model | 119 | -0.17 | -0.43 | 0.096 | 0.2121 |
| Global methylation | Age 9 | European ancestry | prenatal exposures model | 119 | -0.14 | -0.41 | 0.13 | 0.30927 |
| Global methylation | Age 9 | European ancestry | secondhand smoke exposure model | 119 | -0.16 | -0.47 | 0.16 | 0.33736 |
| Global methylation | Age 9 | European ancestry | surrogate variable models | 119 | 0.006 | -0.0096 | 0.022 | 0.44932 |
| Global methylation | Age 9 | Multiethnic sample | base model | 753 | -0.056 | -0.16 | 0.047 | 0.28583 |
| Global methylation | Age 9 | Multiethnic sample | prenatal exposures model | 753 | -0.058 | -0.17 | 0.053 | 0.30745 |
| Global methylation | Age 9 | Multiethnic sample | secondhand smoke exposure model | 753 | -0.07 | -0.2 | 0.063 | 0.30296 |
| Global methylation | Age 9 | Multiethnic sample | surrogate variable models | 753 | 0.0013 | -0.0047 | 0.0073 | 0.67261 |
| Pediatric clock | Age 15 | Admixed ancestry - Latin heritage | base model | 171 | -0.18 | -0.92 | 0.55 | 0.62191 |
| Pediatric clock | Age 15 | Admixed ancestry - Latin heritage | prenatal exposures model | 171 | -0.33 | -1.1 | 0.45 | 0.40657 |
| Pediatric clock | Age 15 | Admixed ancestry - Latin heritage | secondhand smoke exposure model | 171 | -0.99 | -1.8 | -0.13 | 0.024254 |
| Pediatric clock | Age 15 | Admixed ancestry - Latin heritage | surrogate variable models | 171 | 0.3 | -0.17 | 0.78 | 0.20463 |
| Pediatric clock | Age 15 | African ancestry | base model | 457 | -0.042 | -0.41 | 0.33 | 0.82315 |
| Pediatric clock | Age 15 | African ancestry | prenatal exposures model | 457 | -0.057 | -0.46 | 0.35 | 0.78273 |
| Pediatric clock | Age 15 | African ancestry | secondhand smoke exposure model | 457 | -0.13 | -0.63 | 0.37 | 0.60359 |
| Pediatric clock | Age 15 | African ancestry | surrogate variable models | 457 | -0.2 | -0.45 | 0.046 | 0.11082 |
| Pediatric clock | Age 15 | European ancestry | base model | 119 | 0.12 | -0.39 | 0.62 | 0.64348 |
| Pediatric clock | Age 15 | European ancestry | prenatal exposures model | 119 | 0.26 | -0.3 | 0.81 | 0.36188 |
| Pediatric clock | Age 15 | European ancestry | secondhand smoke exposure model | 119 | 0.16 | -0.52 | 0.83 | 0.64938 |
| Pediatric clock | Age 15 | European ancestry | surrogate variable models | 119 | 0.11 | -0.27 | 0.48 | 0.57378 |
| Pediatric clock | Age 15 | Multiethnic sample | base model | 747 | -0.042 | -0.32 | 0.24 | 0.76718 |
| Pediatric clock | Age 15 | Multiethnic sample | prenatal exposures model | 747 | -0.016 | -0.32 | 0.28 | 0.91485 |
| Pediatric clock | Age 15 | Multiethnic sample | secondhand smoke exposure model | 747 | -0.16 | -0.52 | 0.2 | 0.37218 |
| Pediatric clock | Age 15 | Multiethnic sample | surrogate variable models | 747 | -0.11 | -0.29 | 0.076 | 0.25311 |
| Pediatric clock | Age 9 | Admixed ancestry - Latin heritage | base model | 180 | -0.16 | -0.49 | 0.17 | 0.34525 |
| Pediatric clock | Age 9 | Admixed ancestry - Latin heritage | prenatal exposures model | 180 | -0.24 | -0.59 | 0.11 | 0.17781 |
| Pediatric clock | Age 9 | Admixed ancestry - Latin heritage | secondhand smoke exposure model | 180 | -0.33 | -0.74 | 0.081 | 0.11435 |
| Pediatric clock | Age 9 | Admixed ancestry - Latin heritage | surrogate variable models | 180 | 0.14 | -0.17 | 0.45 | 0.36052 |
| Pediatric clock | Age 9 | African ancestry | base model | 454 | -0.098 | -0.31 | 0.11 | 0.36651 |
| Pediatric clock | Age 9 | African ancestry | prenatal exposures model | 454 | -0.14 | -0.37 | 0.092 | 0.2357 |
| Pediatric clock | Age 9 | African ancestry | secondhand smoke exposure model | 454 | -0.18 | -0.46 | 0.1 | 0.20481 |
| Pediatric clock | Age 9 | African ancestry | surrogate variable models | 454 | -0.13 | -0.31 | 0.051 | 0.16087 |
| Pediatric clock | Age 9 | European ancestry | base model | 119 | -0.18 | -0.47 | 0.1 | 0.20782 |
| Pediatric clock | Age 9 | European ancestry | prenatal exposures model | 119 | -0.16 | -0.47 | 0.14 | 0.28562 |
| Pediatric clock | Age 9 | European ancestry | secondhand smoke exposure model | 119 | -0.18 | -0.54 | 0.19 | 0.34053 |
| Pediatric clock | Age 9 | European ancestry | surrogate variable models | 119 | 0.075 | -0.19 | 0.34 | 0.5797 |
| Pediatric clock | Age 9 | Multiethnic sample | base model | 753 | -0.13 | -0.28 | 0.026 | 0.1044 |
| Pediatric clock | Age 9 | Multiethnic sample | prenatal exposures model | 753 | -0.15 | -0.32 | 0.012 | 0.069991 |
| Pediatric clock | Age 9 | Multiethnic sample | secondhand smoke exposure model | 753 | -0.21 | -0.4 | -0.011 | 0.038915 |
| Pediatric clock | Age 9 | Multiethnic sample | surrogate variable models | 753 | -0.065 | -0.19 | 0.064 | 0.3227 |
| 6073 site polymethylation score: coefficients for sustained smoking from Joubert newborn cordblood meta-analysis, w/ cell-type control - PMC4833289 (/n mean-centered (coefficients) & z-score standardized (score)) | Age 15 | Admixed ancestry - Latin heritage | base model | 171 | 0.34 | -0.042 | 0.73 | 0.080637 |
| 6073 site polymethylation score: coefficients for sustained smoking from Joubert newborn cordblood meta-analysis, w/ cell-type control - PMC4833289 (/n mean-centered (coefficients) & z-score standardized (score)) | Age 15 | Admixed ancestry - Latin heritage | prenatal exposures model | 171 | 0.3 | -0.11 | 0.71 | 0.14899 |
| 6073 site polymethylation score: coefficients for sustained smoking from Joubert newborn cordblood meta-analysis, w/ cell-type control - PMC4833289 (/n mean-centered (coefficients) & z-score standardized (score)) | Age 15 | Admixed ancestry - Latin heritage | secondhand smoke exposure model | 171 | 0.28 | -0.19 | 0.75 | 0.24099 |
| 6073 site polymethylation score: coefficients for sustained smoking from Joubert newborn cordblood meta-analysis, w/ cell-type control - PMC4833289 (/n mean-centered (coefficients) & z-score standardized (score)) | Age 15 | Admixed ancestry - Latin heritage | surrogate variable models | 171 | 0.31 | 0.076 | 0.55 | 0.010106 |
| 6073 site polymethylation score: coefficients for sustained smoking from Joubert newborn cordblood meta-analysis, w/ cell-type control - PMC4833289 (/n mean-centered (coefficients) & z-score standardized (score)) | Age 15 | African ancestry | base model | 457 | 0.52 | 0.32 | 0.72 | 3.0662E-07 |
| 6073 site polymethylation score: coefficients for sustained smoking from Joubert newborn cordblood meta-analysis, w/ cell-type control - PMC4833289 (/n mean-centered (coefficients) & z-score standardized (score)) | Age 15 | African ancestry | prenatal exposures model | 457 | 0.55 | 0.33 | 0.76 | 7.2883E-07 |
| 6073 site polymethylation score: coefficients for sustained smoking from Joubert newborn cordblood meta-analysis, w/ cell-type control - PMC4833289 (/n mean-centered (coefficients) & z-score standardized (score)) | Age 15 | African ancestry | secondhand smoke exposure model | 457 | 0.38 | 0.12 | 0.64 | 0.004661 |
| 6073 site polymethylation score: coefficients for sustained smoking from Joubert newborn cordblood meta-analysis, w/ cell-type control - PMC4833289 (/n mean-centered (coefficients) & z-score standardized (score)) | Age 15 | African ancestry | surrogate variable models | 457 | 0.38 | 0.27 | 0.48 | 1.1264E-11 |
| 6073 site polymethylation score: coefficients for sustained smoking from Joubert newborn cordblood meta-analysis, w/ cell-type control - PMC4833289 (/n mean-centered (coefficients) & z-score standardized (score)) | Age 15 | European ancestry | base model | 119 | 0.23 | -0.23 | 0.68 | 0.33156 |
| 6073 site polymethylation score: coefficients for sustained smoking from Joubert newborn cordblood meta-analysis, w/ cell-type control - PMC4833289 (/n mean-centered (coefficients) & z-score standardized (score)) | Age 15 | European ancestry | prenatal exposures model | 119 | 0.39 | -0.11 | 0.89 | 0.12489 |
| 6073 site polymethylation score: coefficients for sustained smoking from Joubert newborn cordblood meta-analysis, w/ cell-type control - PMC4833289 (/n mean-centered (coefficients) & z-score standardized (score)) | Age 15 | European ancestry | secondhand smoke exposure model | 119 | -0.044 | -0.61 | 0.52 | 0.87854 |
| 6073 site polymethylation score: coefficients for sustained smoking from Joubert newborn cordblood meta-analysis, w/ cell-type control - PMC4833289 (/n mean-centered (coefficients) & z-score standardized (score)) | Age 15 | European ancestry | surrogate variable models | 119 | 0.43 | 0.22 | 0.65 | 0.00013998 |
| 6073 site polymethylation score: coefficients for sustained smoking from Joubert newborn cordblood meta-analysis, w/ cell-type control - PMC4833289 (/n mean-centered (coefficients) & z-score standardized (score)) | Age 15 | Multiethnic sample | base model | 747 | 0.48 | 0.32 | 0.63 | 4.4196E-09 |
| 6073 site polymethylation score: coefficients for sustained smoking from Joubert newborn cordblood meta-analysis, w/ cell-type control - PMC4833289 (/n mean-centered (coefficients) & z-score standardized (score)) | Age 15 | Multiethnic sample | prenatal exposures model | 747 | 0.5 | 0.33 | 0.68 | 1.0322E-08 |
| 6073 site polymethylation score: coefficients for sustained smoking from Joubert newborn cordblood meta-analysis, w/ cell-type control - PMC4833289 (/n mean-centered (coefficients) & z-score standardized (score)) | Age 15 | Multiethnic sample | secondhand smoke exposure model | 747 | 0.33 | 0.13 | 0.54 | 0.0012659 |
| 6073 site polymethylation score: coefficients for sustained smoking from Joubert newborn cordblood meta-analysis, w/ cell-type control - PMC4833289 (/n mean-centered (coefficients) & z-score standardized (score)) | Age 15 | Multiethnic sample | surrogate variable models | 747 | 0.36 | 0.28 | 0.45 | 3.4416E-16 |
| 6073 site polymethylation score: coefficients for sustained smoking from Joubert newborn cordblood meta-analysis, w/ cell-type control - PMC4833289 (/n mean-centered (coefficients) & z-score standardized (score)) | Age 9 | Admixed ancestry - Latin heritage | base model | 180 | 0.2 | -0.19 | 0.58 | 0.31341 |
| 6073 site polymethylation score: coefficients for sustained smoking from Joubert newborn cordblood meta-analysis, w/ cell-type control - PMC4833289 (/n mean-centered (coefficients) & z-score standardized (score)) | Age 9 | Admixed ancestry - Latin heritage | prenatal exposures model | 180 | 0.2 | -0.21 | 0.62 | 0.33385 |
| 6073 site polymethylation score: coefficients for sustained smoking from Joubert newborn cordblood meta-analysis, w/ cell-type control - PMC4833289 (/n mean-centered (coefficients) & z-score standardized (score)) | Age 9 | Admixed ancestry - Latin heritage | secondhand smoke exposure model | 180 | 0.18 | -0.31 | 0.67 | 0.46088 |
| 6073 site polymethylation score: coefficients for sustained smoking from Joubert newborn cordblood meta-analysis, w/ cell-type control - PMC4833289 (/n mean-centered (coefficients) & z-score standardized (score)) | Age 9 | Admixed ancestry - Latin heritage | surrogate variable models | 180 | 0.22 | -0.0051 | 0.45 | 0.05538 |
| 6073 site polymethylation score: coefficients for sustained smoking from Joubert newborn cordblood meta-analysis, w/ cell-type control - PMC4833289 (/n mean-centered (coefficients) & z-score standardized (score)) | Age 9 | African ancestry | base model | 454 | 0.54 | 0.33 | 0.74 | 3.0597E-07 |
| 6073 site polymethylation score: coefficients for sustained smoking from Joubert newborn cordblood meta-analysis, w/ cell-type control - PMC4833289 (/n mean-centered (coefficients) & z-score standardized (score)) | Age 9 | African ancestry | prenatal exposures model | 454 | 0.56 | 0.34 | 0.78 | 7.7239E-07 |
| 6073 site polymethylation score: coefficients for sustained smoking from Joubert newborn cordblood meta-analysis, w/ cell-type control - PMC4833289 (/n mean-centered (coefficients) & z-score standardized (score)) | Age 9 | African ancestry | secondhand smoke exposure model | 454 | 0.38 | 0.11 | 0.64 | 0.0052312 |
| 6073 site polymethylation score: coefficients for sustained smoking from Joubert newborn cordblood meta-analysis, w/ cell-type control - PMC4833289 (/n mean-centered (coefficients) & z-score standardized (score)) | Age 9 | African ancestry | surrogate variable models | 454 | 0.35 | 0.25 | 0.45 | 3.9463E-11 |
| 6073 site polymethylation score: coefficients for sustained smoking from Joubert newborn cordblood meta-analysis, w/ cell-type control - PMC4833289 (/n mean-centered (coefficients) & z-score standardized (score)) | Age 9 | European ancestry | base model | 119 | 0.55 | 0.15 | 0.95 | 0.0071483 |
| 6073 site polymethylation score: coefficients for sustained smoking from Joubert newborn cordblood meta-analysis, w/ cell-type control - PMC4833289 (/n mean-centered (coefficients) & z-score standardized (score)) | Age 9 | European ancestry | prenatal exposures model | 119 | 0.65 | 0.24 | 1.1 | 0.0022843 |
| 6073 site polymethylation score: coefficients for sustained smoking from Joubert newborn cordblood meta-analysis, w/ cell-type control - PMC4833289 (/n mean-centered (coefficients) & z-score standardized (score)) | Age 9 | European ancestry | secondhand smoke exposure model | 119 | 0.32 | -0.16 | 0.79 | 0.18848 |
| 6073 site polymethylation score: coefficients for sustained smoking from Joubert newborn cordblood meta-analysis, w/ cell-type control - PMC4833289 (/n mean-centered (coefficients) & z-score standardized (score)) | Age 9 | European ancestry | surrogate variable models | 119 | 0.51 | 0.25 | 0.77 | 0.00019439 |
| 6073 site polymethylation score: coefficients for sustained smoking from Joubert newborn cordblood meta-analysis, w/ cell-type control - PMC4833289 (/n mean-centered (coefficients) & z-score standardized (score)) | Age 9 | Multiethnic sample | base model | 753 | 0.51 | 0.35 | 0.66 | 2.983E-10 |
| 6073 site polymethylation score: coefficients for sustained smoking from Joubert newborn cordblood meta-analysis, w/ cell-type control - PMC4833289 (/n mean-centered (coefficients) & z-score standardized (score)) | Age 9 | Multiethnic sample | prenatal exposures model | 753 | 0.54 | 0.38 | 0.71 | 2.8409E-10 |
| 6073 site polymethylation score: coefficients for sustained smoking from Joubert newborn cordblood meta-analysis, w/ cell-type control - PMC4833289 (/n mean-centered (coefficients) & z-score standardized (score)) | Age 9 | Multiethnic sample | secondhand smoke exposure model | 753 | 0.39 | 0.19 | 0.59 | 0.00014114 |
| 6073 site polymethylation score: coefficients for sustained smoking from Joubert newborn cordblood meta-analysis, w/ cell-type control - PMC4833289 (/n mean-centered (coefficients) & z-score standardized (score)) | Age 9 | Multiethnic sample | surrogate variable models | 753 | 0.36 | 0.28 | 0.45 | 1.2799E-16 |
| 28 site polymethylation score (Reese): coefficients for sustained smoking from newborn cordblood LASSO regression - PMC5381987 (/n mean-centered (coefficients) & z-score standardized (score)) | Age 15 | Admixed ancestry - Latin heritage | base model | 171 | 0.64 | 0.24 | 1 | 0.0019968 |
| 28 site polymethylation score (Reese): coefficients for sustained smoking from newborn cordblood LASSO regression - PMC5381987 (/n mean-centered (coefficients) & z-score standardized (score)) | Age 15 | Admixed ancestry - Latin heritage | prenatal exposures model | 171 | 0.61 | 0.19 | 1 | 0.005252 |
| 28 site polymethylation score (Reese): coefficients for sustained smoking from newborn cordblood LASSO regression - PMC5381987 (/n mean-centered (coefficients) & z-score standardized (score)) | Age 15 | Admixed ancestry - Latin heritage | secondhand smoke exposure model | 171 | 0.55 | 0.061 | 1 | 0.027716 |
| 28 site polymethylation score (Reese): coefficients for sustained smoking from newborn cordblood LASSO regression - PMC5381987 (/n mean-centered (coefficients) & z-score standardized (score)) | Age 15 | Admixed ancestry - Latin heritage | surrogate variable models | 171 | 0.59 | 0.23 | 0.95 | 0.0015116 |
| 28 site polymethylation score (Reese): coefficients for sustained smoking from newborn cordblood LASSO regression - PMC5381987 (/n mean-centered (coefficients) & z-score standardized (score)) | Age 15 | African ancestry | base model | 457 | 0.42 | 0.21 | 0.63 | 8.0075E-05 |
| 28 site polymethylation score (Reese): coefficients for sustained smoking from newborn cordblood LASSO regression - PMC5381987 (/n mean-centered (coefficients) & z-score standardized (score)) | Age 15 | African ancestry | prenatal exposures model | 457 | 0.39 | 0.16 | 0.62 | 0.00076131 |
| 28 site polymethylation score (Reese): coefficients for sustained smoking from newborn cordblood LASSO regression - PMC5381987 (/n mean-centered (coefficients) & z-score standardized (score)) | Age 15 | African ancestry | secondhand smoke exposure model | 457 | 0.16 | -0.11 | 0.44 | 0.24908 |
| 28 site polymethylation score (Reese): coefficients for sustained smoking from newborn cordblood LASSO regression - PMC5381987 (/n mean-centered (coefficients) & z-score standardized (score)) | Age 15 | African ancestry | surrogate variable models | 457 | 0.38 | 0.19 | 0.56 | 8.8416E-05 |
| 28 site polymethylation score (Reese): coefficients for sustained smoking from newborn cordblood LASSO regression - PMC5381987 (/n mean-centered (coefficients) & z-score standardized (score)) | Age 15 | European ancestry | base model | 119 | 0.86 | 0.44 | 1.3 | 9.9436E-05 |
| 28 site polymethylation score (Reese): coefficients for sustained smoking from newborn cordblood LASSO regression - PMC5381987 (/n mean-centered (coefficients) & z-score standardized (score)) | Age 15 | European ancestry | prenatal exposures model | 119 | 0.9 | 0.44 | 1.4 | 0.00018872 |
| 28 site polymethylation score (Reese): coefficients for sustained smoking from newborn cordblood LASSO regression - PMC5381987 (/n mean-centered (coefficients) & z-score standardized (score)) | Age 15 | European ancestry | secondhand smoke exposure model | 119 | 0.3 | -0.21 | 0.8 | 0.24888 |
| 28 site polymethylation score (Reese): coefficients for sustained smoking from newborn cordblood LASSO regression - PMC5381987 (/n mean-centered (coefficients) & z-score standardized (score)) | Age 15 | European ancestry | surrogate variable models | 119 | 0.92 | 0.55 | 1.3 | 3.9386E-06 |
| 28 site polymethylation score (Reese): coefficients for sustained smoking from newborn cordblood LASSO regression - PMC5381987 (/n mean-centered (coefficients) & z-score standardized (score)) | Age 15 | Multiethnic sample | base model | 747 | 0.54 | 0.38 | 0.7 | 1.5743E-10 |
| 28 site polymethylation score (Reese): coefficients for sustained smoking from newborn cordblood LASSO regression - PMC5381987 (/n mean-centered (coefficients) & z-score standardized (score)) | Age 15 | Multiethnic sample | prenatal exposures model | 747 | 0.53 | 0.35 | 0.71 | 7.2011E-09 |
| 28 site polymethylation score (Reese): coefficients for sustained smoking from newborn cordblood LASSO regression - PMC5381987 (/n mean-centered (coefficients) & z-score standardized (score)) | Age 15 | Multiethnic sample | secondhand smoke exposure model | 747 | 0.3 | 0.088 | 0.51 | 0.0055202 |
| 28 site polymethylation score (Reese): coefficients for sustained smoking from newborn cordblood LASSO regression - PMC5381987 (/n mean-centered (coefficients) & z-score standardized (score)) | Age 15 | Multiethnic sample | surrogate variable models | 747 | 0.5 | 0.36 | 0.65 | 1.6263E-11 |
| 28 site polymethylation score (Reese): coefficients for sustained smoking from newborn cordblood LASSO regression - PMC5381987 (/n mean-centered (coefficients) & z-score standardized (score)) | Age 9 | Admixed ancestry - Latin heritage | base model | 180 | 0.71 | 0.35 | 1.1 | 0.00012978 |
| 28 site polymethylation score (Reese): coefficients for sustained smoking from newborn cordblood LASSO regression - PMC5381987 (/n mean-centered (coefficients) & z-score standardized (score)) | Age 9 | Admixed ancestry - Latin heritage | prenatal exposures model | 180 | 0.73 | 0.34 | 1.1 | 0.00025912 |
| 28 site polymethylation score (Reese): coefficients for sustained smoking from newborn cordblood LASSO regression - PMC5381987 (/n mean-centered (coefficients) & z-score standardized (score)) | Age 9 | Admixed ancestry - Latin heritage | secondhand smoke exposure model | 180 | 0.57 | 0.12 | 1 | 0.013014 |
| 28 site polymethylation score (Reese): coefficients for sustained smoking from newborn cordblood LASSO regression - PMC5381987 (/n mean-centered (coefficients) & z-score standardized (score)) | Age 9 | Admixed ancestry - Latin heritage | surrogate variable models | 180 | 0.79 | 0.46 | 1.1 | 8.182E-06 |
| 28 site polymethylation score (Reese): coefficients for sustained smoking from newborn cordblood LASSO regression - PMC5381987 (/n mean-centered (coefficients) & z-score standardized (score)) | Age 9 | African ancestry | base model | 454 | 0.42 | 0.22 | 0.63 | 5.1138E-05 |
| 28 site polymethylation score (Reese): coefficients for sustained smoking from newborn cordblood LASSO regression - PMC5381987 (/n mean-centered (coefficients) & z-score standardized (score)) | Age 9 | African ancestry | prenatal exposures model | 454 | 0.37 | 0.15 | 0.59 | 0.0010148 |
| 28 site polymethylation score (Reese): coefficients for sustained smoking from newborn cordblood LASSO regression - PMC5381987 (/n mean-centered (coefficients) & z-score standardized (score)) | Age 9 | African ancestry | secondhand smoke exposure model | 454 | 0.16 | -0.1 | 0.43 | 0.23298 |
| 28 site polymethylation score (Reese): coefficients for sustained smoking from newborn cordblood LASSO regression - PMC5381987 (/n mean-centered (coefficients) & z-score standardized (score)) | Age 9 | African ancestry | surrogate variable models | 454 | 0.38 | 0.2 | 0.55 | 3.3577E-05 |
| 28 site polymethylation score (Reese): coefficients for sustained smoking from newborn cordblood LASSO regression - PMC5381987 (/n mean-centered (coefficients) & z-score standardized (score)) | Age 9 | European ancestry | base model | 119 | 0.72 | 0.3 | 1.1 | 0.0010212 |
| 28 site polymethylation score (Reese): coefficients for sustained smoking from newborn cordblood LASSO regression - PMC5381987 (/n mean-centered (coefficients) & z-score standardized (score)) | Age 9 | European ancestry | prenatal exposures model | 119 | 0.84 | 0.4 | 1.3 | 0.00026727 |
| 28 site polymethylation score (Reese): coefficients for sustained smoking from newborn cordblood LASSO regression - PMC5381987 (/n mean-centered (coefficients) & z-score standardized (score)) | Age 9 | European ancestry | secondhand smoke exposure model | 119 | 0.43 | -0.072 | 0.94 | 0.09217 |
| 28 site polymethylation score (Reese): coefficients for sustained smoking from newborn cordblood LASSO regression - PMC5381987 (/n mean-centered (coefficients) & z-score standardized (score)) | Age 9 | European ancestry | surrogate variable models | 119 | 0.79 | 0.4 | 1.2 | 0.00014863 |
| 28 site polymethylation score (Reese): coefficients for sustained smoking from newborn cordblood LASSO regression - PMC5381987 (/n mean-centered (coefficients) & z-score standardized (score)) | Age 9 | Multiethnic sample | base model | 753 | 0.53 | 0.37 | 0.69 | 9.1853E-11 |
| 28 site polymethylation score (Reese): coefficients for sustained smoking from newborn cordblood LASSO regression - PMC5381987 (/n mean-centered (coefficients) & z-score standardized (score)) | Age 9 | Multiethnic sample | prenatal exposures model | 753 | 0.51 | 0.34 | 0.68 | 6.4188E-09 |
| 28 site polymethylation score (Reese): coefficients for sustained smoking from newborn cordblood LASSO regression - PMC5381987 (/n mean-centered (coefficients) & z-score standardized (score)) | Age 9 | Multiethnic sample | secondhand smoke exposure model | 753 | 0.29 | 0.086 | 0.49 | 0.0053437 |
| 28 site polymethylation score (Reese): coefficients for sustained smoking from newborn cordblood LASSO regression - PMC5381987 (/n mean-centered (coefficients) & z-score standardized (score)) | Age 9 | Multiethnic sample | surrogate variable models | 753 | 0.52 | 0.38 | 0.66 | 2.5837E-13 |
| 19 site polymethylation score (Richmond): coefficients for sustained smoking from Joubert older children peripheral blood meta-analysis - PMC4833289 (/n mean-centered (coefficients) & z-score standardized (score)) | Age 15 | Admixed ancestry - Latin heritage | base model | 171 | 0.6 | 0.26 | 0.95 | 0.00076316 |
| 19 site polymethylation score (Richmond): coefficients for sustained smoking from Joubert older children peripheral blood meta-analysis - PMC4833289 (/n mean-centered (coefficients) & z-score standardized (score)) | Age 15 | Admixed ancestry - Latin heritage | prenatal exposures model | 171 | 0.49 | 0.13 | 0.86 | 0.0080478 |
| 19 site polymethylation score (Richmond): coefficients for sustained smoking from Joubert older children peripheral blood meta-analysis - PMC4833289 (/n mean-centered (coefficients) & z-score standardized (score)) | Age 15 | Admixed ancestry - Latin heritage | secondhand smoke exposure model | 171 | 0.33 | -0.078 | 0.74 | 0.11173 |
| 19 site polymethylation score (Richmond): coefficients for sustained smoking from Joubert older children peripheral blood meta-analysis - PMC4833289 (/n mean-centered (coefficients) & z-score standardized (score)) | Age 15 | Admixed ancestry - Latin heritage | surrogate variable models | 171 | 0.23 | -0.039 | 0.49 | 0.09442 |
| 19 site polymethylation score (Richmond): coefficients for sustained smoking from Joubert older children peripheral blood meta-analysis - PMC4833289 (/n mean-centered (coefficients) & z-score standardized (score)) | Age 15 | African ancestry | base model | 457 | 0.32 | 0.14 | 0.5 | 0.00072227 |
| 19 site polymethylation score (Richmond): coefficients for sustained smoking from Joubert older children peripheral blood meta-analysis - PMC4833289 (/n mean-centered (coefficients) & z-score standardized (score)) | Age 15 | African ancestry | prenatal exposures model | 457 | 0.29 | 0.085 | 0.49 | 0.0054518 |
| 19 site polymethylation score (Richmond): coefficients for sustained smoking from Joubert older children peripheral blood meta-analysis - PMC4833289 (/n mean-centered (coefficients) & z-score standardized (score)) | Age 15 | African ancestry | secondhand smoke exposure model | 457 | 0.068 | -0.18 | 0.31 | 0.58936 |
| 19 site polymethylation score (Richmond): coefficients for sustained smoking from Joubert older children peripheral blood meta-analysis - PMC4833289 (/n mean-centered (coefficients) & z-score standardized (score)) | Age 15 | African ancestry | surrogate variable models | 457 | 0.3 | 0.17 | 0.43 | 7.646E-06 |
| 19 site polymethylation score (Richmond): coefficients for sustained smoking from Joubert older children peripheral blood meta-analysis - PMC4833289 (/n mean-centered (coefficients) & z-score standardized (score)) | Age 15 | European ancestry | base model | 119 | 0.43 | 0.07 | 0.79 | 0.01981 |
| 19 site polymethylation score (Richmond): coefficients for sustained smoking from Joubert older children peripheral blood meta-analysis - PMC4833289 (/n mean-centered (coefficients) & z-score standardized (score)) | Age 15 | European ancestry | prenatal exposures model | 119 | 0.5 | 0.098 | 0.89 | 0.015123 |
| 19 site polymethylation score (Richmond): coefficients for sustained smoking from Joubert older children peripheral blood meta-analysis - PMC4833289 (/n mean-centered (coefficients) & z-score standardized (score)) | Age 15 | European ancestry | secondhand smoke exposure model | 119 | 0.21 | -0.26 | 0.68 | 0.37802 |
| 19 site polymethylation score (Richmond): coefficients for sustained smoking from Joubert older children peripheral blood meta-analysis - PMC4833289 (/n mean-centered (coefficients) & z-score standardized (score)) | Age 15 | European ancestry | surrogate variable models | 119 | 0.61 | 0.33 | 0.89 | 4.3606E-05 |
| 19 site polymethylation score (Richmond): coefficients for sustained smoking from Joubert older children peripheral blood meta-analysis - PMC4833289 (/n mean-centered (coefficients) & z-score standardized (score)) | Age 15 | Multiethnic sample | base model | 747 | 0.39 | 0.24 | 0.53 | 1.8854E-07 |
| 19 site polymethylation score (Richmond): coefficients for sustained smoking from Joubert older children peripheral blood meta-analysis - PMC4833289 (/n mean-centered (coefficients) & z-score standardized (score)) | Age 15 | Multiethnic sample | prenatal exposures model | 747 | 0.35 | 0.2 | 0.51 | 1.1698E-05 |
| 19 site polymethylation score (Richmond): coefficients for sustained smoking from Joubert older children peripheral blood meta-analysis - PMC4833289 (/n mean-centered (coefficients) & z-score standardized (score)) | Age 15 | Multiethnic sample | secondhand smoke exposure model | 747 | 0.15 | -0.037 | 0.33 | 0.11722 |
| 19 site polymethylation score (Richmond): coefficients for sustained smoking from Joubert older children peripheral blood meta-analysis - PMC4833289 (/n mean-centered (coefficients) & z-score standardized (score)) | Age 15 | Multiethnic sample | surrogate variable models | 747 | 0.33 | 0.23 | 0.43 | 6.8053E-10 |
| 19 site polymethylation score (Richmond): coefficients for sustained smoking from Joubert older children peripheral blood meta-analysis - PMC4833289 (/n mean-centered (coefficients) & z-score standardized (score)) | Age 9 | Admixed ancestry - Latin heritage | base model | 180 | 0.5 | 0.18 | 0.82 | 0.0022595 |
| 19 site polymethylation score (Richmond): coefficients for sustained smoking from Joubert older children peripheral blood meta-analysis - PMC4833289 (/n mean-centered (coefficients) & z-score standardized (score)) | Age 9 | Admixed ancestry - Latin heritage | prenatal exposures model | 180 | 0.42 | 0.077 | 0.75 | 0.016442 |
| 19 site polymethylation score (Richmond): coefficients for sustained smoking from Joubert older children peripheral blood meta-analysis - PMC4833289 (/n mean-centered (coefficients) & z-score standardized (score)) | Age 9 | Admixed ancestry - Latin heritage | secondhand smoke exposure model | 180 | 0.18 | -0.21 | 0.57 | 0.37296 |
| 19 site polymethylation score (Richmond): coefficients for sustained smoking from Joubert older children peripheral blood meta-analysis - PMC4833289 (/n mean-centered (coefficients) & z-score standardized (score)) | Age 9 | Admixed ancestry - Latin heritage | surrogate variable models | 180 | 0.38 | 0.11 | 0.65 | 0.0053493 |
| 19 site polymethylation score (Richmond): coefficients for sustained smoking from Joubert older children peripheral blood meta-analysis - PMC4833289 (/n mean-centered (coefficients) & z-score standardized (score)) | Age 9 | African ancestry | base model | 454 | 0.35 | 0.17 | 0.53 | 0.00013807 |
| 19 site polymethylation score (Richmond): coefficients for sustained smoking from Joubert older children peripheral blood meta-analysis - PMC4833289 (/n mean-centered (coefficients) & z-score standardized (score)) | Age 9 | African ancestry | prenatal exposures model | 454 | 0.3 | 0.11 | 0.5 | 0.0021257 |
| 19 site polymethylation score (Richmond): coefficients for sustained smoking from Joubert older children peripheral blood meta-analysis - PMC4833289 (/n mean-centered (coefficients) & z-score standardized (score)) | Age 9 | African ancestry | secondhand smoke exposure model | 454 | 0.11 | -0.13 | 0.34 | 0.36365 |
| 19 site polymethylation score (Richmond): coefficients for sustained smoking from Joubert older children peripheral blood meta-analysis - PMC4833289 (/n mean-centered (coefficients) & z-score standardized (score)) | Age 9 | African ancestry | surrogate variable models | 454 | 0.29 | 0.15 | 0.42 | 2.8144E-05 |
| 19 site polymethylation score (Richmond): coefficients for sustained smoking from Joubert older children peripheral blood meta-analysis - PMC4833289 (/n mean-centered (coefficients) & z-score standardized (score)) | Age 9 | European ancestry | base model | 119 | 0.57 | 0.2 | 0.94 | 0.0030348 |
| 19 site polymethylation score (Richmond): coefficients for sustained smoking from Joubert older children peripheral blood meta-analysis - PMC4833289 (/n mean-centered (coefficients) & z-score standardized (score)) | Age 9 | European ancestry | prenatal exposures model | 119 | 0.7 | 0.31 | 1.1 | 0.00054447 |
| 19 site polymethylation score (Richmond): coefficients for sustained smoking from Joubert older children peripheral blood meta-analysis - PMC4833289 (/n mean-centered (coefficients) & z-score standardized (score)) | Age 9 | European ancestry | secondhand smoke exposure model | 119 | 0.38 | -0.063 | 0.83 | 0.091152 |
| 19 site polymethylation score (Richmond): coefficients for sustained smoking from Joubert older children peripheral blood meta-analysis - PMC4833289 (/n mean-centered (coefficients) & z-score standardized (score)) | Age 9 | European ancestry | surrogate variable models | 119 | 0.59 | 0.29 | 0.88 | 0.00017358 |
| 19 site polymethylation score (Richmond): coefficients for sustained smoking from Joubert older children peripheral blood meta-analysis - PMC4833289 (/n mean-centered (coefficients) & z-score standardized (score)) | Age 9 | Multiethnic sample | base model | 753 | 0.43 | 0.29 | 0.57 | 1.3471E-09 |
| 19 site polymethylation score (Richmond): coefficients for sustained smoking from Joubert older children peripheral blood meta-analysis - PMC4833289 (/n mean-centered (coefficients) & z-score standardized (score)) | Age 9 | Multiethnic sample | prenatal exposures model | 753 | 0.41 | 0.27 | 0.56 | 5.6275E-08 |
| 19 site polymethylation score (Richmond): coefficients for sustained smoking from Joubert older children peripheral blood meta-analysis - PMC4833289 (/n mean-centered (coefficients) & z-score standardized (score)) | Age 9 | Multiethnic sample | secondhand smoke exposure model | 753 | 0.21 | 0.035 | 0.39 | 0.018907 |
| 19 site polymethylation score (Richmond): coefficients for sustained smoking from Joubert older children peripheral blood meta-analysis - PMC4833289 (/n mean-centered (coefficients) & z-score standardized (score)) | Age 9 | Multiethnic sample | surrogate variable models | 753 | 0.35 | 0.24 | 0.45 | 1.4853E-10 |
| 568 site polymethylation score (Richmond): coefficients for sustained smoking from Joubert newborn cordblood meta-analysis - PMC4833289 (/n mean-centered (coefficients) & z-score standardized (score)) | Age 15 | Admixed ancestry - Latin heritage | base model | 171 | 0.34 | -0.051 | 0.74 | 0.086937 |
| 568 site polymethylation score (Richmond): coefficients for sustained smoking from Joubert newborn cordblood meta-analysis - PMC4833289 (/n mean-centered (coefficients) & z-score standardized (score)) | Age 15 | Admixed ancestry - Latin heritage | prenatal exposures model | 171 | 0.33 | -0.09 | 0.75 | 0.12297 |
| 568 site polymethylation score (Richmond): coefficients for sustained smoking from Joubert newborn cordblood meta-analysis - PMC4833289 (/n mean-centered (coefficients) & z-score standardized (score)) | Age 15 | Admixed ancestry - Latin heritage | secondhand smoke exposure model | 171 | 0.25 | -0.24 | 0.73 | 0.31128 |
| 568 site polymethylation score (Richmond): coefficients for sustained smoking from Joubert newborn cordblood meta-analysis - PMC4833289 (/n mean-centered (coefficients) & z-score standardized (score)) | Age 15 | Admixed ancestry - Latin heritage | surrogate variable models | 171 | 0.41 | 0.11 | 0.72 | 0.0088071 |
| 568 site polymethylation score (Richmond): coefficients for sustained smoking from Joubert newborn cordblood meta-analysis - PMC4833289 (/n mean-centered (coefficients) & z-score standardized (score)) | Age 15 | African ancestry | base model | 457 | 0.6 | 0.4 | 0.8 | 6.3977E-09 |
| 568 site polymethylation score (Richmond): coefficients for sustained smoking from Joubert newborn cordblood meta-analysis - PMC4833289 (/n mean-centered (coefficients) & z-score standardized (score)) | Age 15 | African ancestry | prenatal exposures model | 457 | 0.63 | 0.41 | 0.84 | 2.718E-08 |
| 568 site polymethylation score (Richmond): coefficients for sustained smoking from Joubert newborn cordblood meta-analysis - PMC4833289 (/n mean-centered (coefficients) & z-score standardized (score)) | Age 15 | African ancestry | secondhand smoke exposure model | 457 | 0.41 | 0.15 | 0.68 | 0.0023413 |
| 568 site polymethylation score (Richmond): coefficients for sustained smoking from Joubert newborn cordblood meta-analysis - PMC4833289 (/n mean-centered (coefficients) & z-score standardized (score)) | Age 15 | African ancestry | surrogate variable models | 457 | 0.49 | 0.34 | 0.63 | 6.5684E-11 |
| 568 site polymethylation score (Richmond): coefficients for sustained smoking from Joubert newborn cordblood meta-analysis - PMC4833289 (/n mean-centered (coefficients) & z-score standardized (score)) | Age 15 | European ancestry | base model | 119 | 0.64 | 0.2 | 1.1 | 0.0047247 |
| 568 site polymethylation score (Richmond): coefficients for sustained smoking from Joubert newborn cordblood meta-analysis - PMC4833289 (/n mean-centered (coefficients) & z-score standardized (score)) | Age 15 | European ancestry | prenatal exposures model | 119 | 0.78 | 0.3 | 1.3 | 0.0015834 |
| 568 site polymethylation score (Richmond): coefficients for sustained smoking from Joubert newborn cordblood meta-analysis - PMC4833289 (/n mean-centered (coefficients) & z-score standardized (score)) | Age 15 | European ancestry | secondhand smoke exposure model | 119 | 0.26 | -0.26 | 0.77 | 0.31993 |
| 568 site polymethylation score (Richmond): coefficients for sustained smoking from Joubert newborn cordblood meta-analysis - PMC4833289 (/n mean-centered (coefficients) & z-score standardized (score)) | Age 15 | European ancestry | surrogate variable models | 119 | 0.69 | 0.39 | 0.99 | 1.6424E-05 |
| 568 site polymethylation score (Richmond): coefficients for sustained smoking from Joubert newborn cordblood meta-analysis - PMC4833289 (/n mean-centered (coefficients) & z-score standardized (score)) | Age 15 | Multiethnic sample | base model | 747 | 0.59 | 0.44 | 0.75 | 4.0742E-13 |
| 568 site polymethylation score (Richmond): coefficients for sustained smoking from Joubert newborn cordblood meta-analysis - PMC4833289 (/n mean-centered (coefficients) & z-score standardized (score)) | Age 15 | Multiethnic sample | prenatal exposures model | 747 | 0.63 | 0.46 | 0.8 | 1.2082E-12 |
| 568 site polymethylation score (Richmond): coefficients for sustained smoking from Joubert newborn cordblood meta-analysis - PMC4833289 (/n mean-centered (coefficients) & z-score standardized (score)) | Age 15 | Multiethnic sample | secondhand smoke exposure model | 747 | 0.42 | 0.21 | 0.62 | 5.8337E-05 |
| 568 site polymethylation score (Richmond): coefficients for sustained smoking from Joubert newborn cordblood meta-analysis - PMC4833289 (/n mean-centered (coefficients) & z-score standardized (score)) | Age 15 | Multiethnic sample | surrogate variable models | 747 | 0.5 | 0.39 | 0.61 | 5.4211E-17 |
| 568 site polymethylation score (Richmond): coefficients for sustained smoking from Joubert newborn cordblood meta-analysis - PMC4833289 (/n mean-centered (coefficients) & z-score standardized (score)) | Age 9 | Admixed ancestry - Latin heritage | base model | 180 | 0.45 | 0.056 | 0.84 | 0.025253 |
| 568 site polymethylation score (Richmond): coefficients for sustained smoking from Joubert newborn cordblood meta-analysis - PMC4833289 (/n mean-centered (coefficients) & z-score standardized (score)) | Age 9 | Admixed ancestry - Latin heritage | prenatal exposures model | 180 | 0.45 | 0.031 | 0.87 | 0.035462 |
| 568 site polymethylation score (Richmond): coefficients for sustained smoking from Joubert newborn cordblood meta-analysis - PMC4833289 (/n mean-centered (coefficients) & z-score standardized (score)) | Age 9 | Admixed ancestry - Latin heritage | secondhand smoke exposure model | 180 | 0.31 | -0.19 | 0.8 | 0.22389 |
| 568 site polymethylation score (Richmond): coefficients for sustained smoking from Joubert newborn cordblood meta-analysis - PMC4833289 (/n mean-centered (coefficients) & z-score standardized (score)) | Age 9 | Admixed ancestry - Latin heritage | surrogate variable models | 180 | 0.46 | 0.17 | 0.75 | 0.0020363 |
| 568 site polymethylation score (Richmond): coefficients for sustained smoking from Joubert newborn cordblood meta-analysis - PMC4833289 (/n mean-centered (coefficients) & z-score standardized (score)) | Age 9 | African ancestry | base model | 454 | 0.61 | 0.41 | 0.82 | 1.0515E-08 |
| 568 site polymethylation score (Richmond): coefficients for sustained smoking from Joubert newborn cordblood meta-analysis - PMC4833289 (/n mean-centered (coefficients) & z-score standardized (score)) | Age 9 | African ancestry | prenatal exposures model | 454 | 0.62 | 0.4 | 0.85 | 6.764E-08 |
| 568 site polymethylation score (Richmond): coefficients for sustained smoking from Joubert newborn cordblood meta-analysis - PMC4833289 (/n mean-centered (coefficients) & z-score standardized (score)) | Age 9 | African ancestry | secondhand smoke exposure model | 454 | 0.38 | 0.12 | 0.65 | 0.005201 |
| 568 site polymethylation score (Richmond): coefficients for sustained smoking from Joubert newborn cordblood meta-analysis - PMC4833289 (/n mean-centered (coefficients) & z-score standardized (score)) | Age 9 | African ancestry | surrogate variable models | 454 | 0.45 | 0.31 | 0.6 | 1.8867E-09 |
| 568 site polymethylation score (Richmond): coefficients for sustained smoking from Joubert newborn cordblood meta-analysis - PMC4833289 (/n mean-centered (coefficients) & z-score standardized (score)) | Age 9 | European ancestry | base model | 119 | 0.84 | 0.37 | 1.3 | 0.00065863 |
| 568 site polymethylation score (Richmond): coefficients for sustained smoking from Joubert newborn cordblood meta-analysis - PMC4833289 (/n mean-centered (coefficients) & z-score standardized (score)) | Age 9 | European ancestry | prenatal exposures model | 119 | 0.95 | 0.46 | 1.4 | 0.00022662 |
| 568 site polymethylation score (Richmond): coefficients for sustained smoking from Joubert newborn cordblood meta-analysis - PMC4833289 (/n mean-centered (coefficients) & z-score standardized (score)) | Age 9 | European ancestry | secondhand smoke exposure model | 119 | 0.53 | -0.032 | 1.1 | 0.064285 |
| 568 site polymethylation score (Richmond): coefficients for sustained smoking from Joubert newborn cordblood meta-analysis - PMC4833289 (/n mean-centered (coefficients) & z-score standardized (score)) | Age 9 | European ancestry | surrogate variable models | 119 | 0.71 | 0.33 | 1.1 | 0.0003854 |
| 568 site polymethylation score (Richmond): coefficients for sustained smoking from Joubert newborn cordblood meta-analysis - PMC4833289 (/n mean-centered (coefficients) & z-score standardized (score)) | Age 9 | Multiethnic sample | base model | 753 | 0.64 | 0.48 | 0.81 | 2.3706E-14 |
| 568 site polymethylation score (Richmond): coefficients for sustained smoking from Joubert newborn cordblood meta-analysis - PMC4833289 (/n mean-centered (coefficients) & z-score standardized (score)) | Age 9 | Multiethnic sample | prenatal exposures model | 753 | 0.68 | 0.5 | 0.85 | 6.6813E-14 |
| 568 site polymethylation score (Richmond): coefficients for sustained smoking from Joubert newborn cordblood meta-analysis - PMC4833289 (/n mean-centered (coefficients) & z-score standardized (score)) | Age 9 | Multiethnic sample | secondhand smoke exposure model | 753 | 0.45 | 0.24 | 0.65 | 2.1615E-05 |
| 568 site polymethylation score (Richmond): coefficients for sustained smoking from Joubert newborn cordblood meta-analysis - PMC4833289 (/n mean-centered (coefficients) & z-score standardized (score)) | Age 9 | Multiethnic sample | surrogate variable models | 753 | 0.51 | 0.39 | 0.63 | 8.4433E-17 |
| AHRR: cg05575921 | Age 15 | Admixed ancestry - Latin heritage | base model | 171 | 0.92 | -1.2 | 3 | 0.38907 |
| AHRR: cg05575921 | Age 15 | Admixed ancestry - Latin heritage | prenatal exposures model | 171 | 1.2 | -1 | 3.4 | 0.29632 |
| AHRR: cg05575921 | Age 15 | Admixed ancestry - Latin heritage | secondhand smoke exposure model | 171 | 1.2 | -1.3 | 3.7 | 0.35512 |
| AHRR: cg05575921 | Age 15 | Admixed ancestry - Latin heritage | surrogate variable models | 171 | -0.63 | -2 | 0.71 | 0.35206 |
| AHRR: cg05575921 | Age 15 | African ancestry | base model | 457 | -1.1 | -2.1 | -0.06 | 0.038262 |
| AHRR: cg05575921 | Age 15 | African ancestry | prenatal exposures model | 457 | -0.99 | -2.1 | 0.15 | 0.089886 |
| AHRR: cg05575921 | Age 15 | African ancestry | secondhand smoke exposure model | 457 | -0.52 | -1.9 | 0.88 | 0.46689 |
| AHRR: cg05575921 | Age 15 | African ancestry | surrogate variable models | 457 | -1 | -1.7 | -0.28 | 0.0063955 |
| AHRR: cg05575921 | Age 15 | European ancestry | base model | 119 | -2 | -3.7 | -0.26 | 0.024765 |
| AHRR: cg05575921 | Age 15 | European ancestry | prenatal exposures model | 119 | -1.9 | -3.8 | 0.026 | 0.053156 |
| AHRR: cg05575921 | Age 15 | European ancestry | secondhand smoke exposure model | 119 | -0.7 | -2.9 | 1.5 | 0.52984 |
| AHRR: cg05575921 | Age 15 | European ancestry | surrogate variable models | 119 | -0.9 | -2 | 0.21 | 0.11083 |
| AHRR: cg05575921 | Age 15 | Multiethnic sample | base model | 747 | -0.93 | -1.7 | -0.13 | 0.023316 |
| AHRR: cg05575921 | Age 15 | Multiethnic sample | prenatal exposures model | 747 | -0.8 | -1.7 | 0.071 | 0.071856 |
| AHRR: cg05575921 | Age 15 | Multiethnic sample | secondhand smoke exposure model | 747 | -0.37 | -1.4 | 0.67 | 0.48125 |
| AHRR: cg05575921 | Age 15 | Multiethnic sample | surrogate variable models | 747 | -0.85 | -1.4 | -0.33 | 0.0014732 |
| AHRR: cg05575921 | Age 9 | Admixed ancestry - Latin heritage | base model | 180 | 0.011 | -1.7 | 1.7 | 0.98955 |
| AHRR: cg05575921 | Age 9 | Admixed ancestry - Latin heritage | prenatal exposures model | 180 | -0.22 | -2 | 1.6 | 0.81187 |
| AHRR: cg05575921 | Age 9 | Admixed ancestry - Latin heritage | secondhand smoke exposure model | 180 | -0.51 | -2.6 | 1.6 | 0.6319 |
| AHRR: cg05575921 | Age 9 | Admixed ancestry - Latin heritage | surrogate variable models | 180 | -0.92 | -2.1 | 0.27 | 0.129 |
| AHRR: cg05575921 | Age 9 | African ancestry | base model | 454 | -1.3 | -2.2 | -0.46 | 0.0028596 |
| AHRR: cg05575921 | Age 9 | African ancestry | prenatal exposures model | 454 | -1.1 | -2 | -0.12 | 0.026798 |
| AHRR: cg05575921 | Age 9 | African ancestry | secondhand smoke exposure model | 454 | -1.2 | -2.3 | -0.018 | 0.046515 |
| AHRR: cg05575921 | Age 9 | African ancestry | surrogate variable models | 454 | -1.1 | -1.7 | -0.6 | 3.7881E-05 |
| AHRR: cg05575921 | Age 9 | European ancestry | base model | 119 | -0.85 | -2.5 | 0.76 | 0.29733 |
| AHRR: cg05575921 | Age 9 | European ancestry | prenatal exposures model | 119 | -0.82 | -2.5 | 0.85 | 0.3328 |
| AHRR: cg05575921 | Age 9 | European ancestry | secondhand smoke exposure model | 119 | -0.076 | -2.1 | 1.9 | 0.93942 |
| AHRR: cg05575921 | Age 9 | European ancestry | surrogate variable models | 119 | -1.2 | -2.3 | -0.058 | 0.039402 |
| AHRR: cg05575921 | Age 9 | Multiethnic sample | base model | 753 | -0.93 | -1.6 | -0.25 | 0.0072683 |
| AHRR: cg05575921 | Age 9 | Multiethnic sample | prenatal exposures model | 753 | -0.73 | -1.5 | 0.0012 | 0.050386 |
| AHRR: cg05575921 | Age 9 | Multiethnic sample | secondhand smoke exposure model | 753 | -0.65 | -1.5 | 0.23 | 0.14821 |
| AHRR: cg05575921 | Age 9 | Multiethnic sample | surrogate variable models | 753 | -1 | -1.4 | -0.6 | 2.2003E-06 |
| MYO1G: cg04180046 | Age 15 | Admixed ancestry - Latin heritage | base model | 171 | 7.2 | 2.5 | 12 | 0.0026661 |
| MYO1G: cg04180046 | Age 15 | Admixed ancestry - Latin heritage | prenatal exposures model | 171 | 6 | 1.1 | 11 | 0.017425 |
| MYO1G: cg04180046 | Age 15 | Admixed ancestry - Latin heritage | secondhand smoke exposure model | 171 | 5.1 | -0.51 | 11 | 0.074271 |
| MYO1G: cg04180046 | Age 15 | Admixed ancestry - Latin heritage | surrogate variable models | 171 | 1.6 | -2.3 | 5.4 | 0.42948 |
| MYO1G: cg04180046 | Age 15 | African ancestry | base model | 457 | 4.1 | 1.8 | 6.5 | 0.00062813 |
| MYO1G: cg04180046 | Age 15 | African ancestry | prenatal exposures model | 457 | 3.6 | 1 | 6.2 | 0.0059952 |
| MYO1G: cg04180046 | Age 15 | African ancestry | secondhand smoke exposure model | 457 | 1.3 | -1.9 | 4.4 | 0.4258 |
| MYO1G: cg04180046 | Age 15 | African ancestry | surrogate variable models | 457 | 3.7 | 1.8 | 5.7 | 0.0001638 |
| MYO1G: cg04180046 | Age 15 | European ancestry | base model | 119 | 4.6 | -0.46 | 9.7 | 0.073995 |
| MYO1G: cg04180046 | Age 15 | European ancestry | prenatal exposures model | 119 | 6.5 | 0.94 | 12 | 0.022445 |
| MYO1G: cg04180046 | Age 15 | European ancestry | secondhand smoke exposure model | 119 | 2.4 | -4.1 | 8.9 | 0.46703 |
| MYO1G: cg04180046 | Age 15 | European ancestry | surrogate variable models | 119 | 6.7 | 2.8 | 11 | 0.00092856 |
| MYO1G: cg04180046 | Age 15 | Multiethnic sample | base model | 747 | 4.9 | 3 | 6.8 | 5.4914E-07 |
| MYO1G: cg04180046 | Age 15 | Multiethnic sample | prenatal exposures model | 747 | 4.5 | 2.5 | 6.6 | 1.9124E-05 |
| MYO1G: cg04180046 | Age 15 | Multiethnic sample | secondhand smoke exposure model | 747 | 2.4 | -0.0042 | 4.9 | 0.050395 |
| MYO1G: cg04180046 | Age 15 | Multiethnic sample | surrogate variable models | 747 | 4 | 2.5 | 5.5 | 3.3058E-07 |
| MYO1G: cg04180046 | Age 9 | Admixed ancestry - Latin heritage | base model | 180 | 6.2 | 2.1 | 10 | 0.0030176 |
| MYO1G: cg04180046 | Age 9 | Admixed ancestry - Latin heritage | prenatal exposures model | 180 | 4.6 | 0.28 | 8.9 | 0.036818 |
| MYO1G: cg04180046 | Age 9 | Admixed ancestry - Latin heritage | secondhand smoke exposure model | 180 | 2 | -2.9 | 6.9 | 0.41999 |
| MYO1G: cg04180046 | Age 9 | Admixed ancestry - Latin heritage | surrogate variable models | 180 | 4.7 | 1.1 | 8.2 | 0.01087 |
| MYO1G: cg04180046 | Age 9 | African ancestry | base model | 454 | 4.2 | 1.9 | 6.5 | 0.0003598 |
| MYO1G: cg04180046 | Age 9 | African ancestry | prenatal exposures model | 454 | 3.9 | 1.4 | 6.4 | 0.0025154 |
| MYO1G: cg04180046 | Age 9 | African ancestry | secondhand smoke exposure model | 454 | 1.8 | -1.3 | 4.8 | 0.25262 |
| MYO1G: cg04180046 | Age 9 | African ancestry | surrogate variable models | 454 | 3.3 | 1.3 | 5.2 | 0.001144 |
| MYO1G: cg04180046 | Age 9 | European ancestry | base model | 119 | 6.7 | 1.7 | 12 | 0.0095165 |
| MYO1G: cg04180046 | Age 9 | European ancestry | prenatal exposures model | 119 | 9.1 | 4 | 14 | 0.00061946 |
| MYO1G: cg04180046 | Age 9 | European ancestry | secondhand smoke exposure model | 119 | 5 | -0.9 | 11 | 0.095431 |
| MYO1G: cg04180046 | Age 9 | European ancestry | surrogate variable models | 119 | 7.1 | 3 | 11 | 0.00081876 |
| MYO1G: cg04180046 | Age 9 | Multiethnic sample | base model | 753 | 5.2 | 3.4 | 7 | 1.69E-08 |
| MYO1G: cg04180046 | Age 9 | Multiethnic sample | prenatal exposures model | 753 | 5.1 | 3.1 | 7 | 3.5591E-07 |
| MYO1G: cg04180046 | Age 9 | Multiethnic sample | secondhand smoke exposure model | 753 | 2.8 | 0.46 | 5.1 | 0.01891 |
| MYO1G: cg04180046 | Age 9 | Multiethnic sample | surrogate variable models | 753 | 4.2 | 2.7 | 5.7 | 4.0837E-08 |
| CYP1A1: cg05549655 | Age 15 | Admixed ancestry - Latin heritage | base model | 171 | 4.1 | 1.6 | 6.5 | 0.0012749 |
| CYP1A1: cg05549655 | Age 15 | Admixed ancestry - Latin heritage | prenatal exposures model | 171 | 3.6 | 1 | 6.2 | 0.0062571 |
| CYP1A1: cg05549655 | Age 15 | Admixed ancestry - Latin heritage | secondhand smoke exposure model | 171 | 2.9 | -0.069 | 5.8 | 0.055556 |
| CYP1A1: cg05549655 | Age 15 | Admixed ancestry - Latin heritage | surrogate variable models | 171 | 3.7 | 1.3 | 6 | 0.002404 |
| CYP1A1: cg05549655 | Age 15 | African ancestry | base model | 457 | 2 | 0.63 | 3.3 | 0.0040556 |
| CYP1A1: cg05549655 | Age 15 | African ancestry | prenatal exposures model | 457 | 1.8 | 0.34 | 3.3 | 0.016045 |
| CYP1A1: cg05549655 | Age 15 | African ancestry | secondhand smoke exposure model | 457 | 0.38 | -1.4 | 2.2 | 0.67926 |
| CYP1A1: cg05549655 | Age 15 | African ancestry | surrogate variable models | 457 | 1.8 | 0.76 | 2.9 | 0.00091307 |
| CYP1A1: cg05549655 | Age 15 | European ancestry | base model | 119 | 3.6 | 1.2 | 6.1 | 0.004143 |
| CYP1A1: cg05549655 | Age 15 | European ancestry | prenatal exposures model | 119 | 3.1 | 0.46 | 5.8 | 0.021943 |
| CYP1A1: cg05549655 | Age 15 | European ancestry | secondhand smoke exposure model | 119 | 2.1 | -1.1 | 5.3 | 0.19646 |
| CYP1A1: cg05549655 | Age 15 | European ancestry | surrogate variable models | 119 | 4.6 | 2.5 | 6.8 | 5.0018E-05 |
| CYP1A1: cg05549655 | Age 15 | Multiethnic sample | base model | 747 | 2.7 | 1.6 | 3.7 | 3.693E-07 |
| CYP1A1: cg05549655 | Age 15 | Multiethnic sample | prenatal exposures model | 747 | 2.5 | 1.4 | 3.6 | 1.043E-05 |
| CYP1A1: cg05549655 | Age 15 | Multiethnic sample | secondhand smoke exposure model | 747 | 1.3 | -0.0019 | 2.6 | 0.050332 |
| CYP1A1: cg05549655 | Age 15 | Multiethnic sample | surrogate variable models | 747 | 2.5 | 1.7 | 3.4 | 7.9268E-09 |
| CYP1A1: cg05549655 | Age 9 | Admixed ancestry - Latin heritage | base model | 180 | 3.2 | 0.62 | 5.9 | 0.015691 |
| CYP1A1: cg05549655 | Age 9 | Admixed ancestry - Latin heritage | prenatal exposures model | 180 | 2.7 | -0.14 | 5.5 | 0.06279 |
| CYP1A1: cg05549655 | Age 9 | Admixed ancestry - Latin heritage | secondhand smoke exposure model | 180 | 2.2 | -1.1 | 5.4 | 0.19679 |
| CYP1A1: cg05549655 | Age 9 | Admixed ancestry - Latin heritage | surrogate variable models | 180 | 3.5 | 1.1 | 5.9 | 0.0041877 |
| CYP1A1: cg05549655 | Age 9 | African ancestry | base model | 454 | 2.2 | 0.79 | 3.5 | 0.0020348 |
| CYP1A1: cg05549655 | Age 9 | African ancestry | prenatal exposures model | 454 | 1.8 | 0.32 | 3.3 | 0.017342 |
| CYP1A1: cg05549655 | Age 9 | African ancestry | secondhand smoke exposure model | 454 | 0.24 | -1.5 | 2 | 0.79227 |
| CYP1A1: cg05549655 | Age 9 | African ancestry | surrogate variable models | 454 | 1.7 | 0.55 | 2.8 | 0.0035335 |
| CYP1A1: cg05549655 | Age 9 | European ancestry | base model | 119 | 3.2 | 0.77 | 5.6 | 0.010363 |
| CYP1A1: cg05549655 | Age 9 | European ancestry | prenatal exposures model | 119 | 3.1 | 0.56 | 5.7 | 0.017272 |
| CYP1A1: cg05549655 | Age 9 | European ancestry | secondhand smoke exposure model | 119 | 1.5 | -1.5 | 4.4 | 0.32564 |
| CYP1A1: cg05549655 | Age 9 | European ancestry | surrogate variable models | 119 | 3.6 | 1.4 | 5.8 | 0.0015525 |
| CYP1A1: cg05549655 | Age 9 | Multiethnic sample | base model | 753 | 2.6 | 1.6 | 3.6 | 1.1498E-06 |
| CYP1A1: cg05549655 | Age 9 | Multiethnic sample | prenatal exposures model | 753 | 2.4 | 1.2 | 3.5 | 3.7737E-05 |
| CYP1A1: cg05549655 | Age 9 | Multiethnic sample | secondhand smoke exposure model | 753 | 1.1 | -0.26 | 2.4 | 0.11399 |
| CYP1A1: cg05549655 | Age 9 | Multiethnic sample | surrogate variable models | 753 | 2.1 | 1.2 | 3 | 2.288E-06 |
| GFI1: cg14179389 | Age 15 | Admixed ancestry - Latin heritage | base model | 171 | -0.47 | -1.7 | 0.73 | 0.44162 |
| GFI1: cg14179389 | Age 15 | Admixed ancestry - Latin heritage | prenatal exposures model | 171 | -0.71 | -2 | 0.54 | 0.26242 |
| GFI1: cg14179389 | Age 15 | Admixed ancestry - Latin heritage | secondhand smoke exposure model | 171 | -0.54 | -2 | 0.9 | 0.45883 |
| GFI1: cg14179389 | Age 15 | Admixed ancestry - Latin heritage | surrogate variable models | 171 | -1.1 | -2.3 | 0.11 | 0.075293 |
| GFI1: cg14179389 | Age 15 | African ancestry | base model | 457 | -0.75 | -1.3 | -0.22 | 0.0057657 |
| GFI1: cg14179389 | Age 15 | African ancestry | prenatal exposures model | 457 | -0.68 | -1.3 | -0.1 | 0.0215 |
| GFI1: cg14179389 | Age 15 | African ancestry | secondhand smoke exposure model | 457 | -0.6 | -1.3 | 0.12 | 0.1014 |
| GFI1: cg14179389 | Age 15 | African ancestry | surrogate variable models | 457 | -0.86 | -1.4 | -0.35 | 0.00093844 |
| GFI1: cg14179389 | Age 15 | European ancestry | base model | 119 | -2.2 | -3.9 | -0.54 | 0.010299 |
| GFI1: cg14179389 | Age 15 | European ancestry | prenatal exposures model | 119 | -1.9 | -3.8 | -0.037 | 0.045726 |
| GFI1: cg14179389 | Age 15 | European ancestry | secondhand smoke exposure model | 119 | -2 | -4.2 | 0.31 | 0.089921 |
| GFI1: cg14179389 | Age 15 | European ancestry | surrogate variable models | 119 | -0.93 | -1.9 | 0.074 | 0.068854 |
| GFI1: cg14179389 | Age 15 | Multiethnic sample | base model | 747 | -0.93 | -1.4 | -0.44 | 0.00017237 |
| GFI1: cg14179389 | Age 15 | Multiethnic sample | prenatal exposures model | 747 | -0.89 | -1.4 | -0.37 | 0.00090322 |
| GFI1: cg14179389 | Age 15 | Multiethnic sample | secondhand smoke exposure model | 747 | -0.73 | -1.4 | -0.11 | 0.021679 |
| GFI1: cg14179389 | Age 15 | Multiethnic sample | surrogate variable models | 747 | -0.97 | -1.4 | -0.55 | 6.0301E-06 |
| GFI1: cg14179389 | Age 9 | Admixed ancestry - Latin heritage | base model | 180 | -0.86 | -2.1 | 0.37 | 0.16852 |
| GFI1: cg14179389 | Age 9 | Admixed ancestry - Latin heritage | prenatal exposures model | 180 | -1.1 | -2.4 | 0.2 | 0.09601 |
| GFI1: cg14179389 | Age 9 | Admixed ancestry - Latin heritage | secondhand smoke exposure model | 180 | -0.74 | -2.3 | 0.8 | 0.34209 |
| GFI1: cg14179389 | Age 9 | Admixed ancestry - Latin heritage | surrogate variable models | 180 | -1.3 | -2.5 | -0.018 | 0.046869 |
| GFI1: cg14179389 | Age 9 | African ancestry | base model | 454 | -0.5 | -1.1 | 0.1 | 0.10427 |
| GFI1: cg14179389 | Age 9 | African ancestry | prenatal exposures model | 454 | -0.43 | -1.1 | 0.23 | 0.19938 |
| GFI1: cg14179389 | Age 9 | African ancestry | secondhand smoke exposure model | 454 | -0.039 | -0.83 | 0.76 | 0.92278 |
| GFI1: cg14179389 | Age 9 | African ancestry | surrogate variable models | 454 | -0.7 | -1.3 | -0.13 | 0.015507 |
| GFI1: cg14179389 | Age 9 | European ancestry | base model | 119 | -1.8 | -3.1 | -0.58 | 0.0044616 |
| GFI1: cg14179389 | Age 9 | European ancestry | prenatal exposures model | 119 | -1.5 | -2.8 | -0.23 | 0.02135 |
| GFI1: cg14179389 | Age 9 | European ancestry | secondhand smoke exposure model | 119 | -1.3 | -2.8 | 0.28 | 0.10857 |
| GFI1: cg14179389 | Age 9 | European ancestry | surrogate variable models | 119 | -2.6 | -3.9 | -1.3 | 0.00010329 |
| GFI1: cg14179389 | Age 9 | Multiethnic sample | base model | 753 | -0.77 | -1.3 | -0.29 | 0.0018099 |
| GFI1: cg14179389 | Age 9 | Multiethnic sample | prenatal exposures model | 753 | -0.73 | -1.3 | -0.21 | 0.0059742 |
| GFI1: cg14179389 | Age 9 | Multiethnic sample | secondhand smoke exposure model | 753 | -0.33 | -0.96 | 0.29 | 0.29368 |
| GFI1: cg14179389 | Age 9 | Multiethnic sample | surrogate variable models | 753 | -1.1 | -1.5 | -0.64 | 2.3979E-06 |
| MYO1G: cg22132788 | Age 15 | Admixed ancestry - Latin heritage | base model | 171 | 6 | 1.8 | 10 | 0.0057341 |
| MYO1G: cg22132788 | Age 15 | Admixed ancestry - Latin heritage | prenatal exposures model | 171 | 4.2 | -0.16 | 8.7 | 0.058739 |
| MYO1G: cg22132788 | Age 15 | Admixed ancestry - Latin heritage | secondhand smoke exposure model | 171 | 3.4 | -1.6 | 8.4 | 0.1761 |
| MYO1G: cg22132788 | Age 15 | Admixed ancestry - Latin heritage | surrogate variable models | 171 | 1.6 | -1.8 | 5 | 0.34957 |
| MYO1G: cg22132788 | Age 15 | African ancestry | base model | 457 | 3.4 | 1.3 | 5.4 | 0.0014777 |
| MYO1G: cg22132788 | Age 15 | African ancestry | prenatal exposures model | 457 | 3.1 | 0.89 | 5.4 | 0.0063955 |
| MYO1G: cg22132788 | Age 15 | African ancestry | secondhand smoke exposure model | 457 | 1.2 | -1.5 | 4 | 0.38552 |
| MYO1G: cg22132788 | Age 15 | African ancestry | surrogate variable models | 457 | 2.8 | 1.3 | 4.4 | 0.00032224 |
| MYO1G: cg22132788 | Age 15 | European ancestry | base model | 119 | 3.4 | -0.62 | 7.4 | 0.096067 |
| MYO1G: cg22132788 | Age 15 | European ancestry | prenatal exposures model | 119 | 4.6 | 0.22 | 9 | 0.040028 |
| MYO1G: cg22132788 | Age 15 | European ancestry | secondhand smoke exposure model | 119 | 1.8 | -3.4 | 7 | 0.49597 |
| MYO1G: cg22132788 | Age 15 | European ancestry | surrogate variable models | 119 | 4.7 | 1.6 | 7.8 | 0.0031256 |
| MYO1G: cg22132788 | Age 15 | Multiethnic sample | base model | 747 | 3.6 | 2 | 5.2 | 2.0247E-05 |
| MYO1G: cg22132788 | Age 15 | Multiethnic sample | prenatal exposures model | 747 | 3.2 | 1.4 | 5 | 0.00049155 |
| MYO1G: cg22132788 | Age 15 | Multiethnic sample | secondhand smoke exposure model | 747 | 1.6 | -0.51 | 3.7 | 0.13531 |
| MYO1G: cg22132788 | Age 15 | Multiethnic sample | surrogate variable models | 747 | 2.8 | 1.6 | 4 | 9.156E-06 |
| MYO1G: cg22132788 | Age 9 | Admixed ancestry - Latin heritage | base model | 180 | 6.1 | 2.4 | 9.8 | 0.0013503 |
| MYO1G: cg22132788 | Age 9 | Admixed ancestry - Latin heritage | prenatal exposures model | 180 | 5.5 | 1.6 | 9.5 | 0.006364 |
| MYO1G: cg22132788 | Age 9 | Admixed ancestry - Latin heritage | secondhand smoke exposure model | 180 | 3.9 | -0.6 | 8.4 | 0.088686 |
| MYO1G: cg22132788 | Age 9 | Admixed ancestry - Latin heritage | surrogate variable models | 180 | 3.3 | 0.014 | 6.6 | 0.049072 |
| MYO1G: cg22132788 | Age 9 | African ancestry | base model | 454 | 3.3 | 1.4 | 5.2 | 0.00078449 |
| MYO1G: cg22132788 | Age 9 | African ancestry | prenatal exposures model | 454 | 3 | 0.91 | 5.1 | 0.004979 |
| MYO1G: cg22132788 | Age 9 | African ancestry | secondhand smoke exposure model | 454 | 1.3 | -1.2 | 3.8 | 0.30888 |
| MYO1G: cg22132788 | Age 9 | African ancestry | surrogate variable models | 454 | 2.6 | 1.1 | 4.1 | 0.00054734 |
| MYO1G: cg22132788 | Age 9 | European ancestry | base model | 119 | 3.1 | -0.9 | 7 | 0.12814 |
| MYO1G: cg22132788 | Age 9 | European ancestry | prenatal exposures model | 119 | 4.7 | 0.59 | 8.7 | 0.025383 |
| MYO1G: cg22132788 | Age 9 | European ancestry | secondhand smoke exposure model | 119 | 2.4 | -2.4 | 7.2 | 0.31674 |
| MYO1G: cg22132788 | Age 9 | European ancestry | surrogate variable models | 119 | 2.4 | -0.45 | 5.3 | 0.096941 |
| MYO1G: cg22132788 | Age 9 | Multiethnic sample | base model | 753 | 3.7 | 2.2 | 5.2 | 2.1512E-06 |
| MYO1G: cg22132788 | Age 9 | Multiethnic sample | prenatal exposures model | 753 | 3.6 | 2 | 5.3 | 1.2069E-05 |
| MYO1G: cg22132788 | Age 9 | Multiethnic sample | secondhand smoke exposure model | 753 | 2.2 | 0.29 | 4.1 | 0.024105 |
| MYO1G: cg22132788 | Age 9 | Multiethnic sample | surrogate variable models | 753 | 2.6 | 1.4 | 3.8 | 1.278E-05 |

Supplemental Figure 5 DNA methylation summary measures across combinations of exposure to prenatal and postnatal maternal smoking. Sample size is shown at the bottom of violin plots with captured boxplots. Polymethylation score shown in top facets, percent DNA methylation at cg05575921 in AHRR gene shown in bottom facets, age 9 samples shown in left facets, age 15 shown in right facets

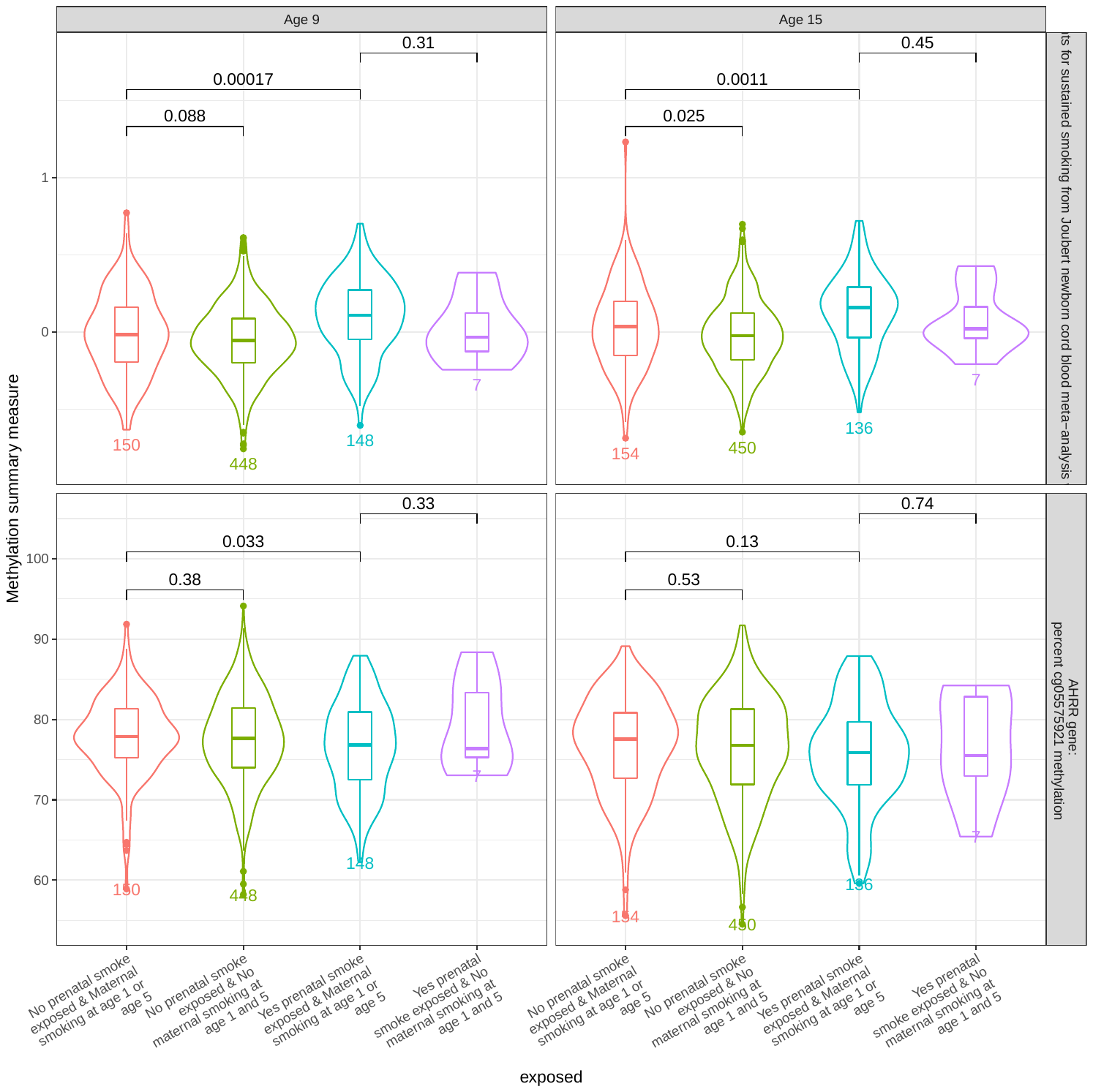

Supplemental Figure 6: Heatmap of *P* values from Pearson correlations between known covariates and all surrogate variables from surrogate variable models calculated using sva function in R

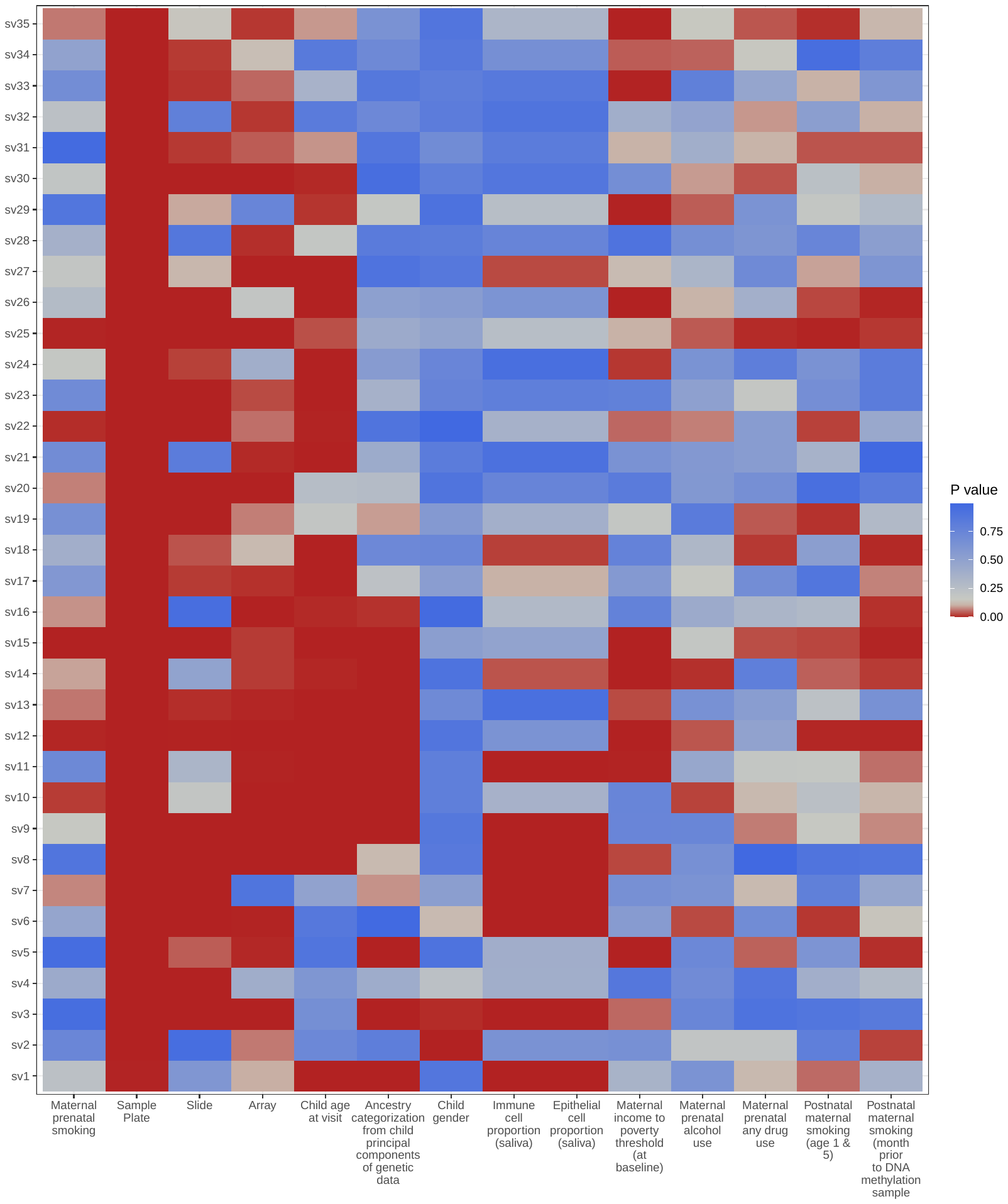

Supplemental Figure 7: Associations between prenatal maternal smoking and polymethylation scores calculated using alternative coefficient sets among 805 children in the Fragile Families and Child Wellbeing study. Base model: Child sex, child age, indicator variable for residence in Detroit, Chicago or Toledo, maternal income to poverty ratio, immune cell proportion, plate, 1st two principal components from PCA on child genetic data, child age. Prenatal exposures model: base model + yes/no any prenatal maternal alcohol use, yes/no any prenatal other drug use. Secondhand smoking models: prenatal exposures model + yes/no any postnatal maternal/primary care giver smoking at age 1 or 5, pack/day smoking in month prior to primary caregiver/maternal interview at age 9 or 15. Surrogate variable: including all surrogate variables

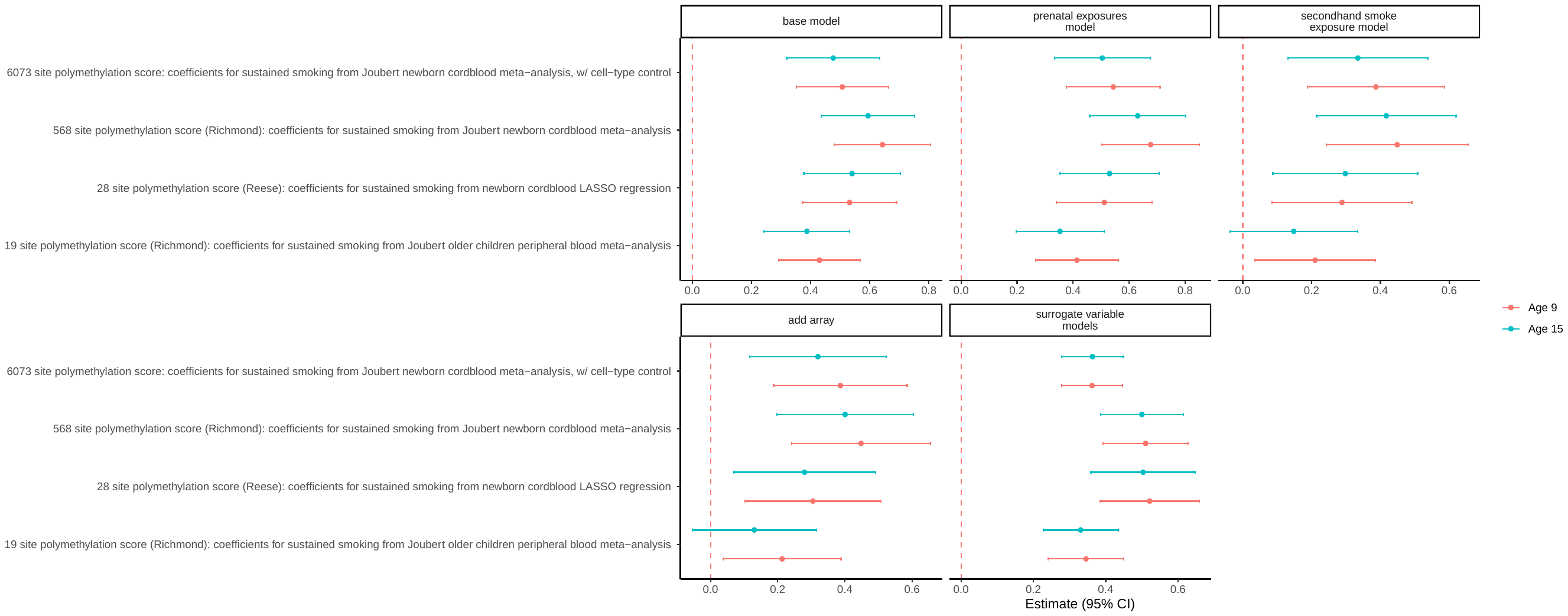

Supplemental Figure 8 - Associations between prenatal maternal smoking and other *a priori* CpG sites selected from previous meta-analyses among 811 children in the Fragile Families and Child Wellbeing study. Fragile Families and Child Wellbeing models controlled for Child sex, maternal income to poverty ratio, immune cell proportion, plate, 1st two principal components from PCA on child genetic data, child age.

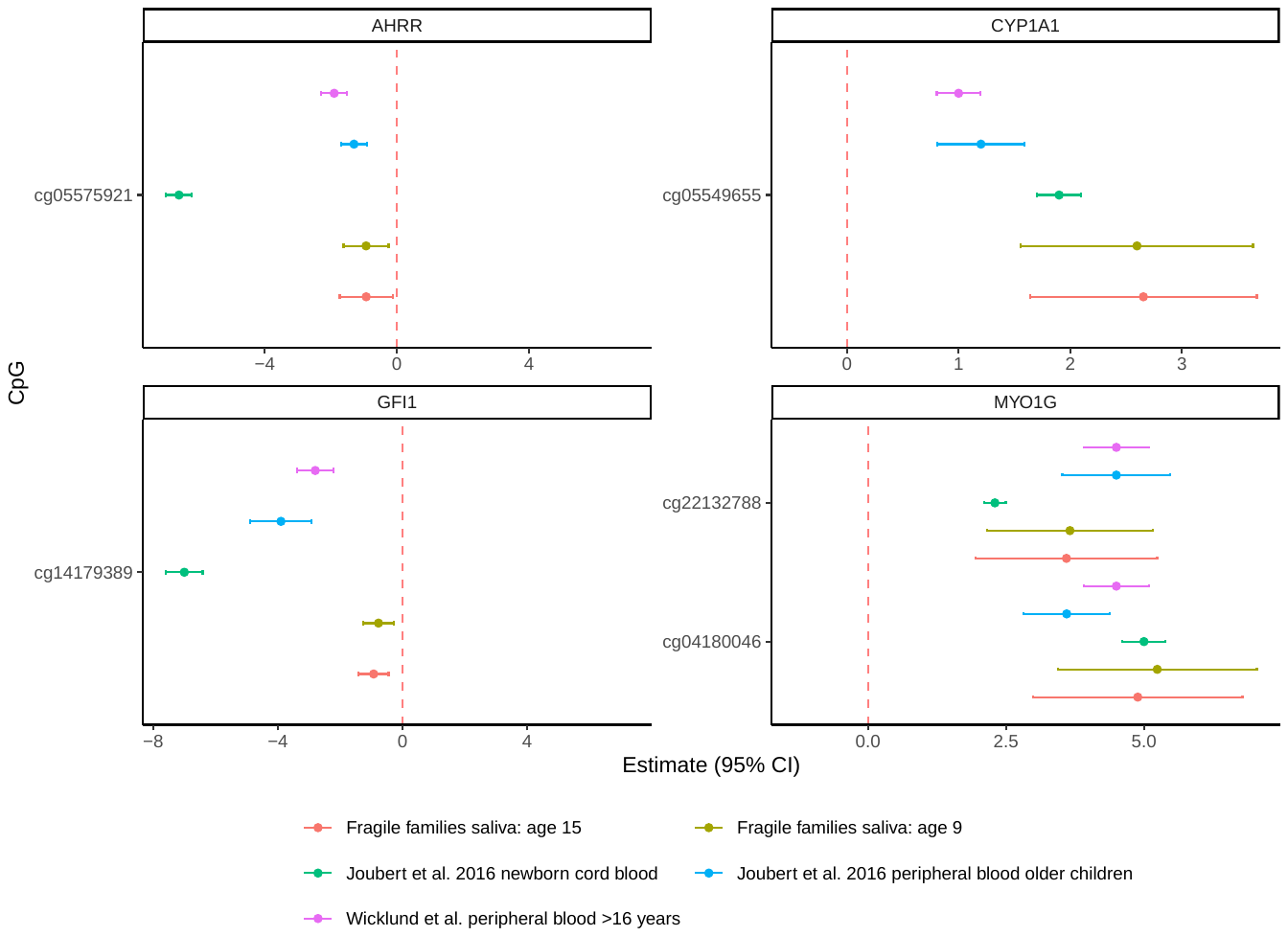

Supplemental Table 5: Area under the curve for models shown in Main Figure 4 for classifying prenatal maternal smoke exposure, with *P* value when comparing to base model only or to model using polymethylation score + base model. Base model: Child sex, maternal income to poverty ratio, indicator variable for residence in Detroit, Toledo or Chicago, immune cell proportion, plate, 1st two principal components from PCA on child genetic data, child age.

| Age strata | Model type | Area under the curve (base model+DNA methylation summary measure) | Area under the curve (base model alone) | Difference (AUC - AUC of base model) (95% CI - DeLong) | P value vs base model (DeLong) |
| --- | --- | --- | --- | --- | --- |
| Age 9 | Base model+Global methylation | 0.73 | 0.73 | 0 (95% CI: (-0.007, 0.005)) | 0.7382 |
| Age 9 | Base model+6073 site polymethylation score: coefficients for sustained smoking from Joubert newborn cordblood meta-analysis, w/ cell-type control - PMC4833289 (/n mean-centered (coefficients) & z-score standardized (score)) | 0.77 | 0.73 | 0.05 (95% CI: (0.021, 0.072)) | 4E-04 |
| Age 9 | Base model+AHRR: cg05575921 | 0.74 | 0.73 | 0.01 (95% CI: (-0.005, 0.026)) | 0.1932 |
| Age 15 | Base model+Global methylation | 0.72 | 0.72 | 0 (95% CI: (-0.001, 0)) | 0.475 |
| Age 15 | Base model+6073 site polymethylation score: coefficients for sustained smoking from Joubert newborn cordblood meta-analysis, w/ cell-type control - PMC4833289 (/n mean-centered (coefficients) & z-score standardized (score)) | 0.76 | 0.72 | 0.05 (95% CI: (0.022, 0.075)) | 4E-04 |
| Age 15 | Base model+AHRR: cg05575921 | 0.72 | 0.72 | 0.01 (95% CI: (-0.008, 0.023)) | 0.336 |

*Poylmethylation score constructed using regression coefficients from a model of prenatal maternal smoking and DNA methylation in newborn cord blood as weights. DNA methylation beta values from Fragile Families and Child Wellbeing study were mean-centered, weighted by the regression coefficients and summed. The resulting scores were z-score standardized and used as outcomes in these regression models

Supplemental Table 6: Area under the curve for polymethylation scores (used alone) and polymethylation scores in conjunction with base model variables. Base model: Child sex, maternal income to poverty ratio, indicator variable for residence in Detroit, Toledo or Chicago, immune cell proportion, plate, 1st two principal components from PCA on child genetic data, child age.

| Age strata | Model | AUC | 95% CI (deLong) |
| --- | --- | --- | --- |
| Age 9 | AHRR: cg05575921 | 0.55 | 0.49, 0.6 |
| Age 9 | Base model+AHRR: cg05575921 | 0.74 | 0.69, 0.78 |
| Age 9 | Elastic net score | 0.68 | 0.63, 0.73 |
| Age 9 | 19 site polymethylation score (Richmond): coefficients for sustained smoking from Joubert older children peripheral blood meta-analysis - PMC4833289 (/n mean-centered (coefficients) & z-score standardized (score)) | 0.64 | 0.59, 0.69 |
| Age 9 | Base model+19 site polymethylation score (Richmond): coefficients for sustained smoking from Joubert older children peripheral blood meta-analysis - PMC4833289 (/n mean-centered (coefficients) & z-score standardized (score)) | 0.76 | 0.72, 0.81 |
| Age 9 | 568 site polymethylation score (Richmond): coefficients for sustained smoking from Joubert newborn cordblood meta-analysis - PMC4833289 (/n mean-centered (coefficients) & z-score standardized (score)) | 0.69 | 0.64, 0.74 |
| Age 9 | Base model+568 site polymethylation score (Richmond): coefficients for sustained smoking from Joubert newborn cordblood meta-analysis - PMC4833289 (/n mean-centered (coefficients) & z-score standardized (score)) | 0.78 | 0.74, 0.83 |
| Age 9 | 6073 site polymethylation score: coefficients for sustained smoking from Joubert newborn cordblood meta-analysis, w/ cell-type control - PMC4833289 (/n mean-centered (coefficients) & z-score standardized (score)) | 0.66 | 0.61, 0.71 |
| Age 9 | Base model+6073 site polymethylation score: coefficients for sustained smoking from Joubert newborn cordblood meta-analysis, w/ cell-type control - PMC4833289 (/n mean-centered (coefficients) & z-score standardized (score)) | 0.77 | 0.73, 0.81 |
| Age 15 | AHRR: cg05575921 | 0.53 | 0.48, 0.59 |
| Age 15 | Base model+AHRR: cg05575921 | 0.72 | 0.68, 0.77 |
| Age 15 | Elastic net score | 0.67 | 0.62, 0.72 |
| Age 15 | 19 site polymethylation score (Richmond): coefficients for sustained smoking from Joubert older children peripheral blood meta-analysis - PMC4833289 (/n mean-centered (coefficients) & z-score standardized (score)) | 0.61 | 0.56, 0.66 |
| Age 15 | Base model+19 site polymethylation score (Richmond): coefficients for sustained smoking from Joubert older children peripheral blood meta-analysis - PMC4833289 (/n mean-centered (coefficients) & z-score standardized (score)) | 0.75 | 0.7, 0.79 |
| Age 15 | 568 site polymethylation score (Richmond): coefficients for sustained smoking from Joubert newborn cordblood meta-analysis - PMC4833289 (/n mean-centered (coefficients) & z-score standardized (score)) | 0.68 | 0.63, 0.73 |
| Age 15 | Base model+568 site polymethylation score (Richmond): coefficients for sustained smoking from Joubert newborn cordblood meta-analysis - PMC4833289 (/n mean-centered (coefficients) & z-score standardized (score)) | 0.77 | 0.73, 0.81 |
| Age 15 | 6073 site polymethylation score: coefficients for sustained smoking from Joubert newborn cordblood meta-analysis, w/ cell-type control - PMC4833289 (/n mean-centered (coefficients) & z-score standardized (score)) | 0.66 | 0.61, 0.71 |
| Age 15 | Base model+6073 site polymethylation score: coefficients for sustained smoking from Joubert newborn cordblood meta-analysis, w/ cell-type control - PMC4833289 (/n mean-centered (coefficients) & z-score standardized (score)) | 0.76 | 0.72, 0.81 |
